# Virtual spatial transcriptomics links kidney histopathology to disease-associated molecular programs

**DOI:** 10.64898/2026.11.17.26368273

**Authors:** Kalin Nonchev, Yunsu Jang, Stellor Nlandu Khodo, Viktor H. Koelzer, Gunnar Rätsch, Matthias B. Moor

**Author notes:** **Correspondence:** Kalin Nonchev, Department of Computer Science, ETH Zurich, Universitätstrasse 6, 8092 Zurich, Switzerland. or Matthias B. Moor, Department of Nephrology, University Hospital Bern, Freiburgstrasse 15, 3010 Bern, Switzerland. V.H. Koelzer, G. Rätsch, and M.B. Moor contributed equally as shared last authors.

## Abstract

**Introduction:** Spatial transcriptomics can resolve compartment-specific molecular programs in kidney disease, but its cost and technical requirements limit its use in clinical diagnostics and in large biopsy archives. Predicting spatial gene expression from routine histology with artificial intelligence could extend molecular characterization to existing clinically annotated kidney tissue collections.

**Methods:** We adapted DeepSpot-M, a pretrained multimodal foundation model that predicts spatial gene expression from histology, using paired hematoxylin and eosin images and Visium spatial transcriptomics from 209 Kidney Precision Medicine Project (KPMP) participants. We evaluated virtual profiles in held-out human biopsies, associated them with histopathologic and biochemical injury markers in 567 rats with defined nephrotoxin exposures from a public toxicogenomics database, and generated a virtual atlas from 1 534 histology slides of 476 additional KPMP participants.

**Results:** Kidney-specific adaptation of DeepSpot-M doubled the mean Pearson correlation of each model’s 1 000 best-predicted genes (0.43 versus 0.22) and exceeded recent foundation models. Virtual profiles preserved the direction of all 13 disease-marker differences in measured expression, including tubular injury, maladaptive repair, gluconeogenesis, and fibrosis-associated programs. All 11 measured associations between molecular programs and estimated glomerular filtration rate (eGFR) were also preserved. Without rat-specific training, a virtual injury score distinguished moderate or severe histopathologic lesions (area under the curve, 0.85) and at least twofold blood urea nitrogen elevation (0.82). In our atlas of 5.46 million virtual spots, higher proximal tubule program scores were associated with higher eGFR (partial *r*, 0.32) and less interstitial fibrosis (−0.57), and program–pathology associations agreed with those of measured Visium profiles in independent participants (Spearman *ρ*, 0.64).

**Conclusions:** Kidney-adapted virtual spatial transcriptomics links histology-derived molecular programs to experimental renal injury and human clinical and histologic phenotypes. Our approach enables spatial molecular analyses in routine biopsy histology, which could help decipher disease-associated pathways in existing biopsy archives.

**Translational Statement:** Spatial molecular profiling of kidney biopsies is informative but too costly for routine use, whereas hematoxylin and eosin sections exist for nearly every biopsy. We show that a model adapted to paired kidney histology and spatial transcriptomics infers programs of tubular injury, metabolic differentiation, and fibrosis from routine histology, and that these programs track kidney function and pathology across 476 participants. Applied to existing archives, virtual spatial transcriptomics could stratify heterogeneous patients by tissue molecular state, prioritize biopsies for targeted assays, and support biomarker discovery and trial enrichment once validated prospectively.

## Introduction

Acute kidney injury (AKI) and chronic kidney disease (CKD) shift the balance between differentiated nephron function, epithelial repair, and interstitial remodeling. Single-cell and spatial transcriptomic studies, among them those of the Kidney Precision Medicine Project (KPMP), resolved injury and repair within specific nephron segments and tissue neighborhoods.^1–3^ In mice and humans, proximal tubular cells either regained differentiated functions after injury or persisted in altered states with inflammatory and profibrotic programs,^2,4^ and spatial studies located these injured epithelial populations alongside immune cells, fibroblasts, and myofibroblasts.^5–8^ Such unresolved injury states were associated with functional decline in the human kidney,^3^ indicating that gene expression programs carry information beyond the morphological appearance of the tissue. Among these programs, *HAVCR1* (kidney injury molecule-1) marked acutely injured proximal tubules, *VCAM1* marked tubular cells that failed to repair,^2,4^ and *MMP7*, a Wnt/*β*-catenin target, linked epithelial injury to experimental fibrosis.^9^ In contrast, reduced expression of the gluconeogenic genes *PC*, *PCK1*, *FBP1*, and *G6PC1* marked loss of differentiated proximal tubular function in CKD.^10^ Jointly characterizing these partly opposing programs within the same tissue can help distinguish preserved tubular function from injury, inflammation, and fibrosis.

However, the high cost, low throughput, and specialized tissue requirements of spatial transcriptomic assays confine them to research centers and preclude their use in large kidney cohorts and routine diagnostics. As a consequence, few or no molecular data exist for patients with rarer kidney diseases. By contrast, routine hematoxylin and eosin (H&E)-stained sections are widely available in biopsy archives and are often linked to pathology and clinical information, but they provide no direct measurement of gene expression. Computational pathology has automated the segmentation of renal structures, the detection and grading of lesions, and the mapping of immune cells on these images,^11–19^ yet these approaches quantify tissue morphology without measuring the underlying molecular programs.

Predicting gene expression directly from histology, termed virtual spatial transcriptomics, moves this approach from assigning predefined labels to inferring spatial molecular profiles across the tissue.^20–23^ Recent foundation models, deep learning models pretrained on large spatial transcriptomics collections, predicted thousands of genes from H&E images,^24–26^ and such virtual profiles uncovered molecular biomarkers in routine breast cancer pathology.^27^ These models, however, were developed predominantly in oncology. It therefore remains unresolved whether virtual spatial transcriptomics in the kidney merely reproduces morphology-correlated gene expression or truly recovers renal disease programs and their links to kidney injury, function, and pathology.

To address this gap, we adapted DeepSpot-M, a multimodal foundation model that predicts gene expression from histology,^24^ to paired kidney images and spatial transcriptomic measurements of KPMP (Figure 1). DeepSpot-M represents each gene by its protein sequence.^28,29^ Therefore, the same model can be applied across species without species-specific training. In held-out human biopsies, we compared virtual with measured gene expression, including disease-associated molecular differences and associations with estimated glomerular filtration rate (eGFR). In public data from rats with defined nephrotoxic exposures, we related virtual injury programs to the site and severity of histopathologic injury and to blood urea nitrogen (BUN), thus testing the model against injury of known origin.^30^ Finally, we applied the model to 1 534 archived slides of 476 independent KPMP participants to build a virtual spatial transcriptomics atlas that links diagnoses, histopathology, and kidney function to molecular programs. Our aim was to evaluate routine histology as a proxy for spatial molecular profiling.

**Figure 1.**
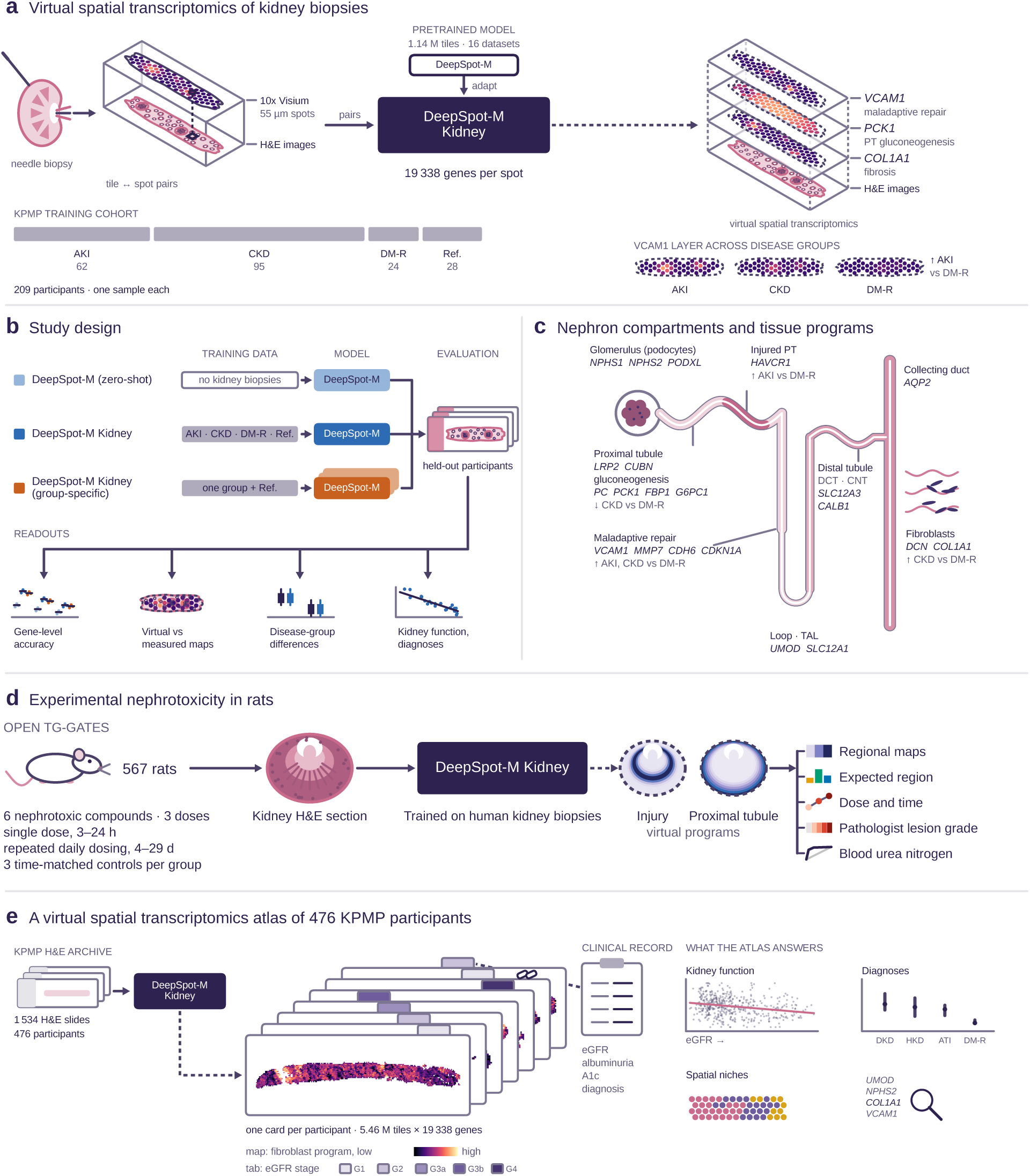
Study design. (a) Hematoxylin and eosin (H&E) tiles centered on 55 µm 10x Genomics Visium spots of Kidney Precision Medicine Project (KPMP) biopsies, paired with the measured expression of each spot, trained DeepSpot-M Kidney (virtual expression of 19 338 genes per spot). Bottom, participants by enrollment group. (b) Model configurations and readouts, evaluated on the same held-out participants. (c) Nephron compartments and tissue programs with marker genes and the expected change relative to DM-R. (d) Kidney H&E sections of Open TG-GATEs rats given nephrotoxic compounds, profiled with DeepSpot-M Kidney without rat-specific training (Figure 3). (e) Virtual atlas of 1 534 KPMP slides from 476 participants without Visium data. AKI, acute kidney injury; ATI, acute tubular injury; CKD, chronic kidney disease; CNT, connecting tubule; DCT, distal convoluted tubule; DKD, diabetic kidney disease; DM-R, diabetes mellitus resilient (diabetes without clinically overt kidney disease); HKD, hypertensive kidney disease; PT, proximal tubule; Ref., reference; TAL, thick ascending limb.

## Methods

### Study population and data

We analyzed paired H&E images and 10x Genomics Visium profiles from 209 KPMP participants, with one frozen specimen per participant embedded in optimal cutting temperature compound (Table 1; Supplementary Table S1).^1,31^ Participants were grouped by KPMP enrollment category into AKI (*n* = 62), CKD (*n* = 95), diabetes mellitus resilient (DM-R, *n* = 24), defined by KPMP as long-standing diabetes without clinically overt kidney disease, and healthy reference tissue (*n* = 28). Clinical information came from the KPMP open-access participant table (Supplementary Methods). KPMP calculated baseline eGFR with the CKD-EPI creatinine equation, for AKI from creatinine values before the injury episode. Therefore, analyses of eGFR excluded participants with AKI. All participants provided informed consent under KPMP protocols approved by the participating institutions’ review boards.

**Table 1.** Characteristics of the 209 Kidney Precision Medicine Project participants by enrollment group.

| Characteristic | AKI (*n*=62) | CKD (*n*=95) | DM-R (*n*=24) | Reference (*n*=28) | All (*n*=209) |
|---|---|---|---|---|---|
| Specimen, *n* (%) |  |  |  |  |  |
| Percutaneous needle biopsy | 61 (98) | 95 (100) | 24 (100) | 0 (0) | 180 (86) |
| Living-donor preperfusion biopsy | 0 (0) | 0 (0) | 0 (0) | 21 (75) | 21 (10) |
| Nephrectomy | 0 (0) | 0 (0) | 0 (0) | 5 (18) | 5 (2) |
| Intraoperative biopsy | 1 (2) | 0 (0) | 0 (0) | 2 (7) | 3 (1) |
| Age, yr, *n* (%) |  |  |  |  |  |
| <40 | 21 (34) | 7 (7) | 6 (25) | 10 (36) | 44 (21) |
| 40–59 | 27 (44) | 32 (34) | 11 (46) | 17 (61) | 87 (42) |
| ≥60 | 14 (23) | 56 (59) | 7 (29) | 1 (4) | 78 (37) |
| Female sex, *n* (%) | 18 (29) | 45 (47) | 15 (63) | 17 (61) | 95 (45) |
| Race, *n* (%)<sup>c</sup> |  |  |  |  |  |
| Asian | 3 (5) | 5 (5) | 0 (0) | 3 (11) | 11 (5) |
| Black or African American | 20 (32) | 29 (31) | 0 (0) | 0 (0) | 49 (23) |
| White | 34 (55) | 50 (53) | 24 (100) | 21 (75) | 129 (62) |
| Other, multiple or not reported | 5 (8) | 11 (12) | 0 (0) | 4 (14) | 20 (10) |
| Baseline eGFR, ml/min per 1.73 m<sup>2</sup>, median (IQR)<sup>a</sup> | 77 (75–92) | 48 (38–58) | 109 (99–114) | 104 (92–111) | 75 (48–97) |
| Participants with a value, *n* | 62 | 94 | 24 | 22 | 202 |
| Diabetes, *n* (%) | 17 (27) | 76 (80) | 24 (100) | 0 (0) | 117 (56) |
| Hypertension, *n* (%) | 30 (48) | 87 (92) | 14 (58) | 2 (7) | 133 (64) |
| RAAS blockade, *n* (%)<sup>b</sup> | 14 (23) | 81 (85) | 12 (50) | – | 107 (51) |
| Albuminuria, *n* (%) |  |  |  |  |  |
| <30 mg/g | 3 (5) | 12 (13) | 23 (96) | 17 (61) | 55 (26) |
| 30 to <300 mg/g | 8 (13) | 23 (24) | 1 (4) | 2 (7) | 34 (16) |
| ≥300 mg/g | 7 (11) | 31 (33) | 0 (0) | 1 (4) | 39 (19) |
| Not recorded | 44 (71) | 29 (31) | 0 (0) | 8 (29) | 81 (39) |
| KDIGO AKI stage, *n* (%) |  |  |  |  |  |
| Stage 1 | 8 (13) | – | – | – | 8 (4) |
| Stage 2 | 10 (16) | – | – | – | 10 (5) |
| Stage 3 | 36 (58) | – | – | – | 36 (17) |
| Not recorded | 8 (13) | – | – | – | 8 (4) |
| Adjudicated primary diagnosis, *n* (%)<sup>d</sup> |  |  |  |  |  |
| Acute tubular injury | 32 (52) | 0 (0) | 0 (0) | 0 (0) | 32 (15) |
| Acute interstitial nephritis | 12 (19) | 0 (0) | 0 (0) | 0 (0) | 12 (6) |
| Diabetic kidney disease | 3 (5) | 42 (44) | 6 (25) | 0 (0) | 51 (24) |
| Hypertensive kidney disease | 0 (0) | 27 (28) | 2 (8) | 0 (0) | 29 (14) |
| No histologic abnormality | 0 (0) | 0 (0) | 8 (33) | 0 (0) | 8 (4) |
| Other | 3 (5) | 12 (13) | 2 (8) | 0 (0) | 17 (8) |
| Not determined or not adjudicated | 12 (19) | 14 (15) | 6 (25) | 28 (100) | 60 (29) |

Values are *n* (%) unless otherwise indicated; percentages are of all participants in the group; –, not recorded or not applicable. AKI, acute kidney injury; CKD, chronic kidney disease; DM-R, diabetes mellitus resilient (long-standing diabetes without clinically overt kidney disease); eGFR, estimated glomerular filtration rate; IQR, interquartile range; KDIGO, Kidney Disease: Improving Global Outcomes; RAAS, renin-angiotensin-aldosterone system.
<sup>a</sup>Baseline eGFR from the KPMP open-access table; for AKI, the value before the injury episode.
<sup>b</sup>Not recorded for reference participants or one participant with CKD; percentages use all participants in each group.
<sup>c</sup>Categories as recorded in the KPMP open-access table. Other, multiple or not reported comprises Other (*n* = 13), more than one category (*n* = 2), and no entry, don’t know, or declined to answer (*n* = 5).
<sup>d</sup>KPMP enrollment groups follow clinical criteria at enrollment, whereas the primary diagnosis was adjudicated afterwards on the biopsy; the two can differ, so that DM-R participants enrolled without clinically overt kidney disease may carry a histologic diagnosis such as diabetic or hypertensive kidney disease.

### Model and kidney adaptation

DeepSpot-M combines histologic features from the Midnight image encoder^32^ with ProtT5 gene embeddings derived from protein sequences^28,29^ through a cross-attention decoder and was developed on 1.14 million spot profiles from 16 cancer spatial transcriptomics datasets without KPMP data.^24^ For kidney adaptation, the image encoder and gene embeddings remained frozen, and the model was trained on H&E tiles centered on the 55 µm Visium spots (65 µm field of view) paired with measured spot profiles (counts per 10 000, log1p transformed; 19 338 genes; Supplementary Methods). We compared the model adapted on pooled kidney biopsies (DeepSpot-M Kidney), models adapted for each enrollment group together with the reference biopsies (DeepSpot-M Kidney (group-specific)), and the pretrained model without kidney adaptation (DeepSpot-M (zero-shot)). For the rat study and the atlas, DeepSpot-M Kidney was adapted on all 209 paired biopsies and applied to whole sections on a lattice of non-overlapping 56 µm tiles, termed virtual spots, each profiled from the 65 µm image crop centered on it.

### Study design and accuracy

Five group-stratified splits at the participant level each used 147 biopsies for adaptation and held out the other 62 for evaluation, and 171 of 209 participants (82%) were held out at least once (Supplementary Figure S1). To reduce the effect of gene dropout,^33,34^ the reference expression of each spot averaged its measured expression with that of its five nearest neighbors in expression space within the same biopsy,^24^ and individual-spot expression served as a sensitivity analysis. Accuracy was the Pearson correlation (*r*) between virtual and reference expression across the spots of a held-out biopsy, averaged over the held-out biopsies of a group. Top-*K* accuracy, the mean *r* of each model’s *K* best-predicted genes ranked by held-out accuracy, is reported as mean±standard deviation (SD) over splits together with a chance level after permuting spots within each held-out biopsy. Gene sets of nephron compartments and tissue programs (Supplementary Table S2) were summarized by the mean *r* of their genes.

Each model family was evaluated before and after fine-tuning on the training biopsies of each split, with the published training objectives for the recent foundation models STPath^25^ and SQUALL^26^ and the kidney adaptation above for DeepSpot-M (Supplementary Methods). As released, STPath and SQUALL had seen 21 KPMP specimens in pretraining,^2^ 19 of which were among the 171 held-out participants. These 19 participants were excluded from all top-*K* comparisons.

### Disease groups and kidney function

We compared AKI and CKD with DM-R biopsies for prespecified injury and maladaptive repair markers (*VCAM1*, *HAVCR1*, *MMP7*, *CDH6*, and *CDKN1A*), gluconeogenic enzymes (*PC*, *PCK1*, *FBP1*, and *G6PC1*), and a fibroblast program score.^2,10^ Pseudo-bulk expression, the mean virtual or measured expression of each gene over the spots of a biopsy, was *z*-scored within each split across the held-out AKI, CKD, and DM-R biopsies, and group differences in mean *z*-score were averaged over splits. A measured difference of at least 0.5 SD of these *z*-scores was counted as reproduced when the virtual difference had the same sign.

Tissue program scores were the mean *z*-score of program genes from published KPMP and kidney atlas studies (Supplementary Table S2),^2,3,35–38^ computed only with models that had not seen the participant and averaged over splits. Program scores were related to eGFR by Spearman correlation in participants with CKD, DM-R, or reference tissue (*n* = 111), and adjudicated diagnoses were compared within AKI (acute interstitial nephritis versus acute tubular injury) and CKD (diabetic versus hypertensive kidney disease) by the difference in mean program score. A measured association whose 95% confidence interval (CI; 10 000 bootstrap resamples of biopsies) excluded zero counted as reproduced when the CI of the virtual association excluded zero on the same side.

### Rat toxicology study

We analyzed kidney H&E sections, histopathologic grades, BUN, and kidney expression arrays of 567 male Sprague-Dawley rats in the public Open TG-GATEs database,^30,39,40^ which received one of six nephrotoxic compounds (cisplatin, carboplatin, gentamicin, 2-bromoethylamine, puromycin aminonucleoside, and doxorubicin) at three doses, or vehicle, for up to eight exposure durations (Supplementary Methods). DeepSpot-M Kidney queried 20 368 rat genes through their ProtT5 embeddings from the STRING database,^29,41^ without rat-specific training or mapping to human orthologs. An injury program (*Lcn2*, *Clu*, *Spp1*, *Timp1*, *Cd44*, and *Sox9*) and a proximal tubule program (*Lrp2*, *Slc34a1*, and *Slc5a2*) were scored in the cortex, outer medulla, and inner medulla with papilla, delimited by depth borders calibrated on the control sections. Program scores averaged, over their genes, the difference in regional mean virtual expression from each rat’s three time-matched controls divided by the pooled control SD, with proximal tubule loss as the negative proximal tubule score. The region of expected injury of each compound was specified from the literature before the analysis. Expression arrays from separate kidney fragments of the same rats were normalized by robust multi-array average^42^ and scored in the same way. Rat data came from a published study whose protocols had been approved by the responsible animal ethics committees.^30^

### KPMP virtual spatial transcriptomics atlas

For the atlas, DeepSpot-M Kidney profiled an independent KPMP histology archive without participant overlap, comprising 1 534 slides from 476 participants, of which 968 were formalin-fixed paraffin-embedded (FFPE) and 566 frozen (Supplementary Methods).

Program scores were computed from the virtual pseudo-bulk of each slide relative to expression-matched control genes and averaged per participant and tissue preparation. Associations with baseline eGFR (participants without AKI), binned albuminuria, proteinuria, and glycated hemoglobin, and pathologist descriptors of the KPMP core (255 participants) were estimated by rank partial correlation adjusted for site, sex, age, and scanner (and cortex fraction for descriptors), separately for FFPE and frozen slides. Partial correlations computed in the same way from measured pseudo-bulk profiles of the 209 Visium participants served as a reference.

Tissue niches were identified on FFPE slides of the Aperio GT450 scanner (583 slides from 293 participants) by Leiden clustering of the virtual expression of program and highly variable genes after Harmony integration across instruments,^43^ annotated by their virtual marker genes and representative H&E tiles, and compared with cell types measured by Xenium in the same participants (Supplementary Methods). To extend Xenium panels, DeepSpot-M Kidney was further adapted to the measured panel genes of single cells in 49 KPMP Xenium sections. The panel genes on chromosomes 1, 3, 5, 7, and 9 were withheld from adaptation, together with their paralogs on other chromosomes, to prevent leakage through the similar protein sequence embeddings of paralogs.^24^ Agreement of the withheld genes with measured expression was compared before and after adaptation by a two-sided Wilcoxon signed-rank test.

### Statistical analysis

In rats with lesions, the region of expected injury was compared with the other two regions, and each region with the matched controls, by two-sided Wilcoxon signed-rank tests, and trends with the duration of dosing were assessed by Spearman correlation. The Benjamini–Hochberg false discovery rate (FDR) was computed over the three compound-specific comparisons of the expected region and controlled at 0.05 over the atlas associations. Areas under the receiver operating characteristic curve (AUCs) were computed with 95% CIs from stratified bootstrap resampling. Two-sided *P* < 0.05 was considered significant. Analyses of disease groups and kidney function in the Visium cohort were descriptive and not corrected for multiple comparisons.

## Results

### Participant characteristics

In the 209 participants with paired histology and Visium profiles, median baseline eGFR ranged from 48 ml/min per 1.73 m^2^ in CKD to 109 ml/min per 1.73 m^2^ in DM-R (Table 1).

### Accuracy of virtual spatial expression in held-out participants

Kidney-specific adaptation improved the agreement between virtual and measured Visium expression in held-out participants, with a mean Pearson correlation of 0.43 for the 1 000 best-predicted genes of DeepSpot-M Kidney, twice that of DeepSpot-M (zero-shot) (0.22). DeepSpot-M Kidney also outperformed the recent foundation models STPath and SQUALL in every disease group, both as released (0.35 and 0.11) and after fine-tuning on the same training biopsies (0.38 and 0.18; Figure 2a). The gain held against individual-spot expression (0.20 versus 0.08; Supplementary Table S3), and the ranking of the models was unchanged with Spearman correlation and for genes ranked by spatial autocorrelation or variability in the training biopsies instead of held-out accuracy, except that fine-tuned STPath was more accurate for the 100 most variable genes (Supplementary Tables S4–S6). DeepSpot-M Kidney performed as well as the group-specific models overall and better on AKI biopsies (Supplementary Figure S2a), and accuracy averaged over all genes improved in 168 of 171 held-out biopsies (Supplementary Figure S3). Furthermore, each doubling of training biopsies increased the gain in accuracy over DeepSpot-M (zero-shot) by 10 (top-100 genes) to 21 percentage points (all genes), without saturation at 147 biopsies (Supplementary Figure S4).

**Figure 2.**
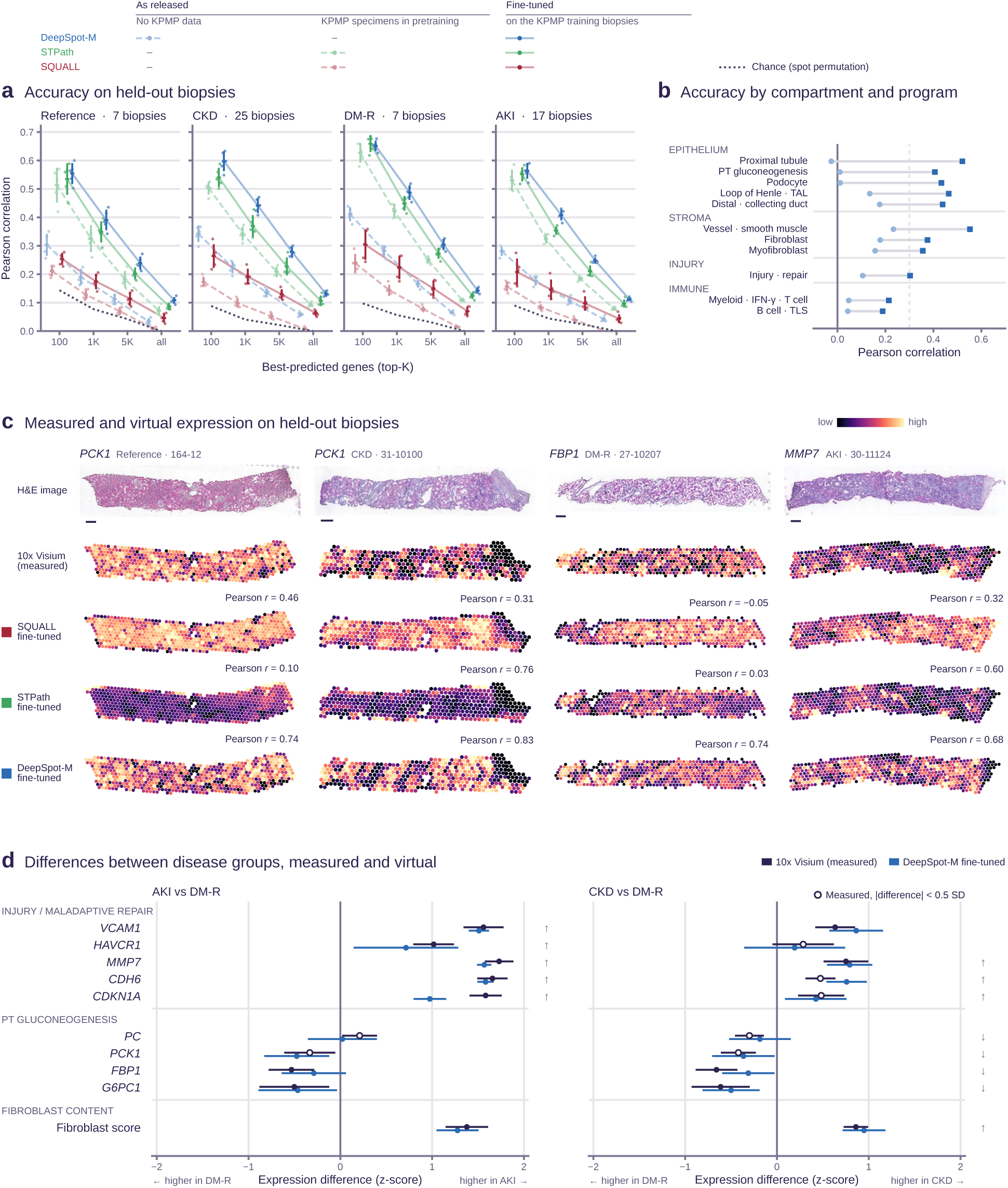
Spatial gene expression and disease-group differences recovered by DeepSpot-M Kidney in held-out participants. (a) Mean Pearson correlation between virtual and reference expression (Methods) of each model’s *K* best-predicted genes on held-out biopsies per group, excluding the 19 held-out participants in the training data of SQUALL and STPath; numbers above the panels, median held-out biopsies per split. Key, model family by training and exposure to KPMP data (DeepSpot-M as released, DeepSpot-M (zero-shot); fine-tuned, DeepSpot-M Kidney); pale dashed lines, as released; full color solid lines, fine-tuned on the training biopsies of each split; dots, the five participant-level splits; horizontal line, mean; vertical line, SD; dotted line, chance (virtual spots permuted within each biopsy). (b) Mean gene-wise correlation within compartment and program gene sets; dashed line, *r* = 0.3. (c) H&E image and measured and virtual maps of one marker on one held-out biopsy per group; numbers, *r*; one color scale per map (2nd–98th percentile). Scale bars, 250 µm. (d) Difference in pseudo-bulk expression (*z*-score; Methods) between AKI or CKD and DM-R; points, mean over five splits; open measured points, differences below 0.5 SD; bars, SD; arrows, prespecified expected direction. AKI, acute kidney injury; CKD, chronic kidney disease; DM-R, diabetes mellitus resilient; H&E, hematoxylin and eosin; IFN, interferon; KPMP, Kidney Precision Medicine Project; PT, proximal tubule; SD, standard deviation; TAL, thick ascending limb; TLS, tertiary lymphoid structure.

Across renal compartments and tissue programs, accuracy was highest for epithelial and fibrosis-associated programs, including proximal tubule, podocyte, vessel, and fibroblast programs (mean *r*, 0.36–0.55), followed by injury and repair (0.30) and immune programs (0.19–0.21; Figure 2b; per-gene accuracy in Supplementary Table S7). When held-out spots were assigned to their highest-scoring program, canonical cell type markers outside the program gene sets peaked in their own compartment for 33 of 41 genes in virtual and 36 in measured expression (Supplementary Figure S5). Spatial maps of *MMP7* in AKI, *PCK1* in CKD and reference tissue, and *FBP1* in DM-R agreed with measured Visium patterns (*r* = 0.68–0.83; Figure 2c).

### Disease-group differences and kidney function associations in virtual profiles

In measured pseudo-bulk expression, prespecified injury and repair markers and the fibroblast program score were higher in AKI and CKD than in DM-R biopsies, whereas gluconeogenic genes were lower, except *PC* in AKI (Figure 2d). Virtual profiles from DeepSpot-M Kidney preserved the direction of all 13 measured differences of at least 0.5 SD (filled points in Figure 2d), and virtual and measured differences were strongly correlated (*r* = 0.97, compared with 0.80 for DeepSpot-M (zero-shot)). DeepSpot-M Kidney also reproduced their size (median ratio of virtual to measured, 0.93), whereas the group-specific models, each applied to biopsies of its own enrollment group, enlarged them (1.54; Supplementary Figure S2b). Therefore, a single model trained across disease groups derives group differences from the tissue alone and can be applied to biopsies of unknown diagnosis, as in the atlas. Deconvolution with the KPMP single-cell atlas^2^ showed lower proximal tubule and higher adaptive proximal tubule and fibroblast fractions in AKI and CKD than in DM-R biopsies in both measured and virtual profiles (Supplementary Figure S6).

Among participants with CKD, DM-R, or reference tissue, the virtual fibroblast program score correlated with eGFR similarly to the measured score (Spearman *ρ*, −0.51 [95% CI, −0.65 to −0.35] versus −0.49 [95% CI, −0.63 to −0.32]; Supplementary Figure S7). Overall, virtual profiles reproduced all 11 program–eGFR associations whose measured CI excluded zero and all eight such differences between diabetic and hypertensive kidney disease. In contrast, the three differences between acute interstitial nephritis and acute tubular injury kept their direction without meeting this criterion.

### Virtual molecular programs in experimental nephrotoxicity

To relate virtual programs to the site and severity of injury of known origin, which human biopsies rarely provide, we applied DeepSpot-M Kidney without rat-specific training to archived kidney sections of 567 rats that had received nephrotoxic compounds under controlled conditions, from the public Open TG-GATEs database (Figure 1d). In rats given cisplatin, virtual injury exceeded that of time-matched controls in the cortex and outer medulla, which contain the proximal straight tubules that cisplatin injures,^44^ but not in the inner medulla (*P* < 0.001, *P* = 0.006, and *P* = 0.17), and was higher in the outer medulla, the region of expected injury, than in the other two regions (*P* = 0.02; FDR, 0.051). Likewise, injury after 2-bromoethylamine was higher in its region of expected injury, the inner medulla (*P* = 0.04; FDR, 0.054),^45^ whereas proximal tubule loss after gentamicin was not higher in the cortex (*P* = 0.33; FDR, 0.33; Figure 3a and b).^46^ Scores in the region of expected injury increased with the duration of daily dosing for cisplatin injury and for proximal tubule loss after puromycin aminonucleoside and gentamicin (Spearman *ρ*, 0.62, 0.86, and 0.67), whereas after 4 days they did not exceed those of matched controls (Figure 3c; Supplementary Figure S8).

**Figure 3.**
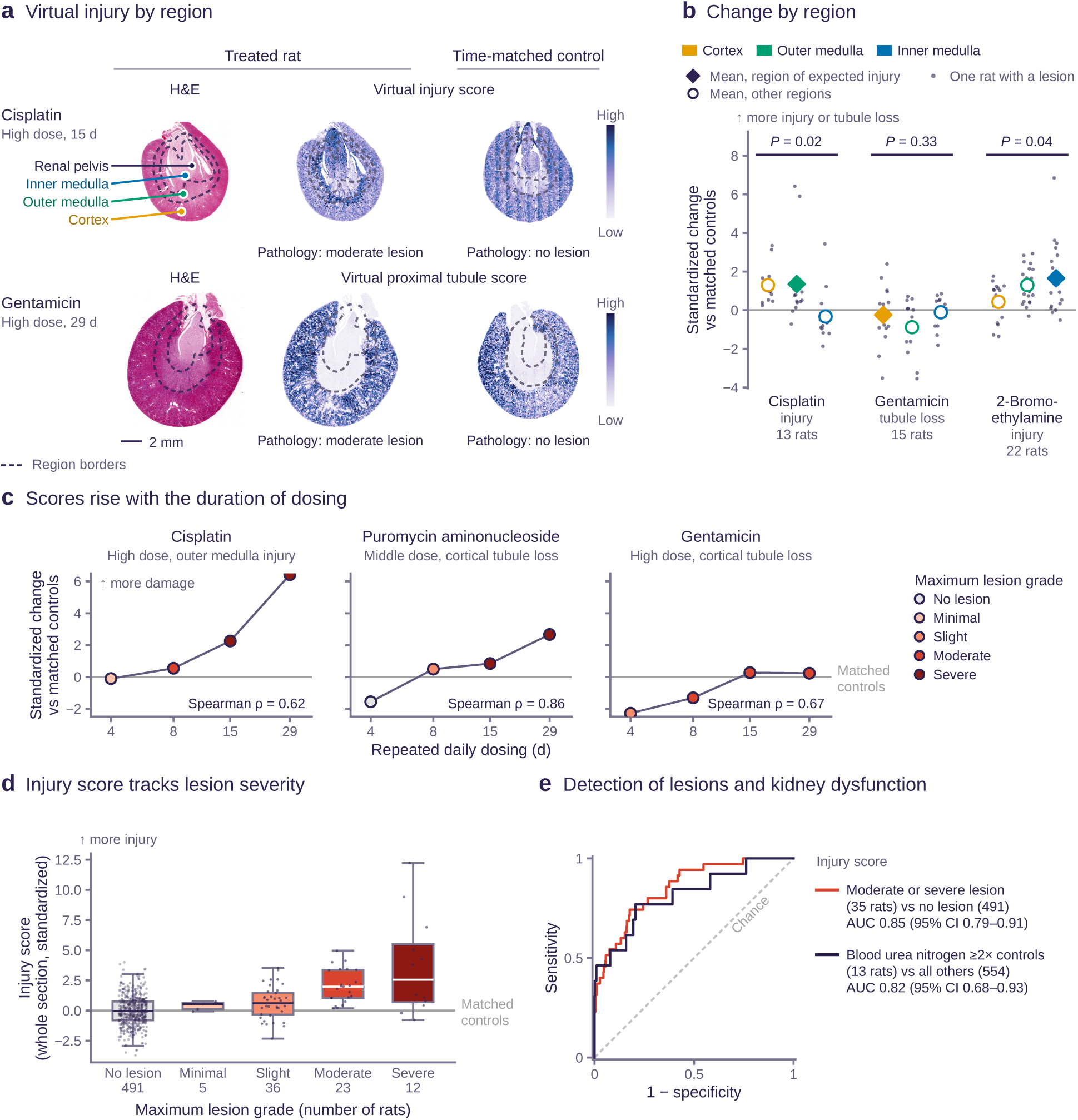
Virtual molecular programs and histopathologic and biochemical injury in experi-mental nephrotoxicity. Kidney sections of 567 Open TG-GATEs rats profiled with DeepSpot-M Kidney (Figure 1d). Scores are standardized against each rat’s three time-matched controls (0, control mean; 1, one control SD). (a) H&E images and virtual injury (cisplatin) and proximal tubule (gentamicin) program maps of a treated rat and its time-matched control; dashed lines, region borders (Methods). Scale bar, 2 mm. (b) Region scores in rats with lesions; small dots, rats; diamonds, mean of the region of expected injury (Methods); open circles, means of the other two regions; *P*, two-sided Wilcoxon signed-rank test, expected versus other regions (nominal; FDR in the text). (c) Score in the region of expected injury over daily dosing, colored by maximum lesion grade; *ρ*, Spearman correlation with time; the 29-day cisplatin point is one surviving rat. (d) Whole-section injury score by maximum lesion grade (boxes, median and interquartile range). (e) Receiver operating characteristic curves of the injury score for moderate or severe lesions versus none (35 versus 491 rats) and for blood urea nitrogen at least twice that of controls (13 versus 554). AUC, area under the curve; CI, confidence interval; H&E, hematoxylin and eosin; SD, standard deviation.

Virtual scores were also associated with independently assessed kidney injury, as the whole-section injury score increased with lesion grade and distinguished moderate or severe lesions from no lesion (AUC, 0.85) and at least twofold BUN elevation relative to matched controls (AUC, 0.82; Figure 3d and e). In animals with lesions, the virtual injury program correlated with the injury program measured by expression arrays in separate kidney fragments (Spearman *ρ*, 0.55 [95% CI, 0.36 to 0.69]), whereas the proximal tubule loss program did not (*ρ*, 0.24 [95% CI, −0.05 to 0.50]), and neither program correlated in animals without lesions (Supplementary Figure S9).

### Tissue niches, clinicopathologic associations, and Xenium validation of the virtual atlas

The atlas comprised 5.46 million virtual spots with expression profiles of 19 338 genes (Figure 4a; Supplementary Figure S10) and was linked to baseline eGFR in 440 participants and to pathologist descriptors in 255. To our knowledge, it is the largest histology-derived virtual spatial transcriptomics atlas of non-neoplastic human kidney disease reported to date.

**Figure 4.**
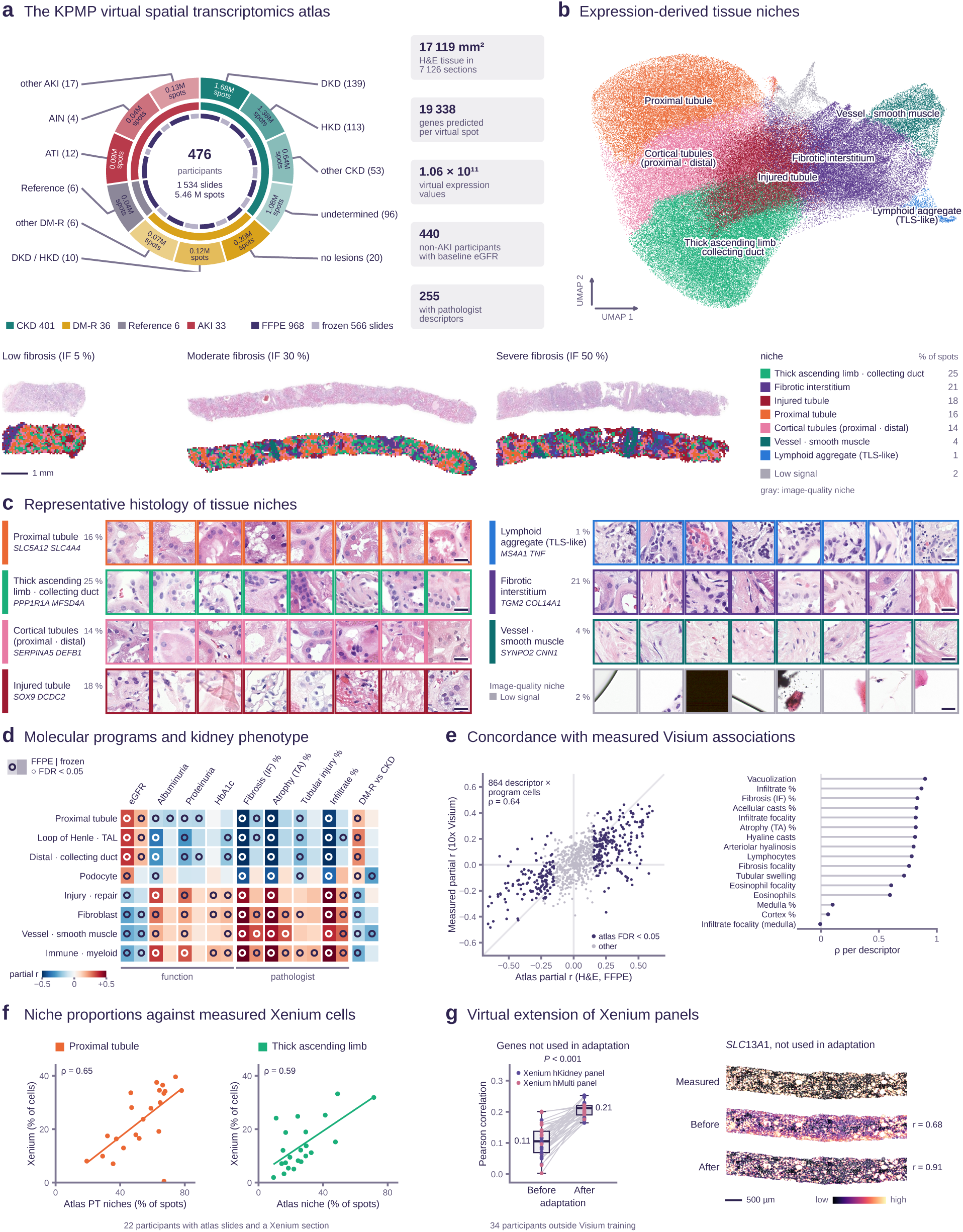
A virtual spatial transcriptomics atlas of the KPMP H&E archive. DeepSpot-M Kidney, adapted on all 209 paired biopsies, was applied to 1 534 open-access KPMP slides from 476 participants without Visium data. (a) Participants by diagnosis and enrollment group, slides by tissue preparation, and virtual spots; right, atlas totals. (b) UMAP of 174 900 virtual spots from 583 FFPE slides of one scanner model; bottom, H&E images and niche maps of three example DKD biopsies with increasing interstitial fibrosis, chosen to illustrate the niches; right, share of spots per niche (rounded percentages). Scale bar, 1 mm. (c) Representative 65 µm H&E input tiles of eight participants per niche (most typical left) and the top two virtual marker genes. Scale bars, 20 µm. H&E images in b and c are adjusted uniformly for display. (d) Partial correlation of participant-level program scores with kidney function and pathologist descriptors (Methods) in FFPE (left half of each cell) and frozen slides of the same participants (right half); circles, FDR < 0.05. (e) Atlas (FFPE) versus measured Visium partial correlations of 864 descriptor–program pairs in other participants (left) and their Spearman correlation for the 16 descriptors with the most atlas associations (right). (f) Atlas niche share versus measured Xenium cell share in the 22 participants with a Xenium section and FFPE slides of the scanner model in b (different tissue pieces); left, proximal tubule, cortical tubule, and injured tubule niches pooled versus proximal tubule cells (including injured); right, thick ascending limb and collecting duct niche versus thick ascending limb cells; dots, participants; line, least-squares fit; *ρ*, Spearman correlation. (g) Xenium panel genes withheld from the adaptation of DeepSpot-M Kidney to the remaining panel genes in the cells of all 49 Xenium sections (Methods). Left, median Pearson *r* per section between virtual and reference expression across cells for the 233 withheld genes on chromosomes 1, 3, 5, 7, and 9 in 34 sections of participants outside the Visium training; dots and lines, sections, colored by Xenium panel; boxes, median and interquartile range; whiskers, 1.5 times the interquartile range; *P*, two-sided Wilcoxon signed-rank test. Right, measured and virtual expression of the proximal tubule sulfate transporter *SLC13A1* in the cells of one section before and after adaptation; numbers, whole-section *r*; one color scale per map (2nd–98th percentile). Scale bar, 500 µm. AIN, acute interstitial nephritis; AKI, acute kidney injury; ATI, acute tubular injury; CKD, chronic kidney disease; DKD, diabetic kidney disease; DM-R, diabetes mellitus resilient; FDR, false discovery rate; FFPE, formalin-fixed paraffin-embedded; H&E, hematoxylin and eosin; HKD, hypertensive kidney disease; IF, interstitial fibrosis; KPMP, Kidney Precision Medicine Project; TA, tubular atrophy; TAL, thick ascending limb; TLS, tertiary lymphoid structure; UMAP, uniform manifold approximation and projection.

Clustering of virtual expression from 583 FFPE slides of one scanner model identified seven tissue niches and one image-quality niche (Figure 4b and c; Supplementary Figure S11a). Their top virtual marker genes matched the histology, for example, *SLC5A12* and *SLC4A4* in proximal tubule and *MS4A1* in lymphoid aggregates resembling tertiary lymphoid structures (Figure 4c). Across 102 participants, proximal tubule regions decreased whereas injured tubule, fibrotic, and lymphoid aggregate regions increased with interstitial fibrosis (Spearman *ρ*, −0.63, 0.47, 0.71, and 0.58; Supplementary Figure S11b).

At the participant level (Figure 4d), higher proximal tubule program scores were associated with higher eGFR (partial *r*, 0.32), less interstitial fibrosis (−0.57), and less tubular atrophy (−0.56). In the 231 FFPE participants with interstitial fibrosis and tubular atrophy scores, the association with eGFR persisted after additional adjustment for both and for diagnosis (partial *r*, 0.18). In cross-validation, adding the virtual program scores to demographic and technical covariates, interstitial fibrosis, and tubular atrophy increased the explained variance of eGFR from 0.35 to 0.41 (Supplementary Table S8). Overall, 64 of 72 program–phenotype associations reached FDR < 0.05 in FFPE tissue, and 54 of these had the same direction in frozen sections from the same participants.

Atlas program–pathology associations agreed with those derived from measured Visium profiles of independent participants across 864 descriptor–program pairs (Spearman *ρ*, 0.64), and strongly for interstitial fibrosis, tubular atrophy, and mononuclear infiltrate (*ρ*, 0.82–0.87; Figure 4e). Beyond predefined programs, exploratory gene-level analyses identified 1 534 genes associated with eGFR at FDR < 0.05 in both tissue preparations with the same direction, including the proximal tubule transporter genes *SLC4A4* and *SLC3A1*, which were higher at higher eGFR, and the extracellular matrix genes *COL14A1* and *OGN*, which were lower (Supplementary Figure S11c).

In the 22 participants with both a Xenium section and niche-annotated FFPE slides, atlas niche shares followed the measured Xenium cell shares (Spearman *ρ*, 0.65 for the proximal tubule niches and 0.59 for thick ascending limb and collecting duct; Figure 4f; Supplementary Figure S12a and b). On the Xenium sections themselves, the share of spots in the virtual injured tubule niche followed the measured share of injured proximal tubule cells (Spearman *ρ*, 0.53; Supplementary Figure S12c and d).

Finally, after adaptation of DeepSpot-M Kidney to the remaining panel genes in single cells of all 49 KPMP Xenium sections, the 233 withheld panel genes agreed more closely with their measured expression (median Pearson *r* per section, 0.11 before and 0.21 after adaptation), with improvement in all 34 sections from participants outside the Visium training (*P* < 0.001; Figure 4g) and gains in every enrollment group, on both Xenium panels, and in the 56 µm spots of the atlas (Supplementary Table S9). Virtual expression of 19 338 genes for the cells of all 49 Xenium sections, extending their 480-gene panels, is provided as a resource (Data Sharing Statement).

## Discussion

In this study, kidney adaptation of DeepSpot-M yielded virtual molecular profiles from standard histology that reproduced nephron compartments, disease-group differences, and associations with kidney function in held-out participants. Beyond the paired cohort, virtual programs tracked graded injury and BUN elevation in rats under controlled nephrotoxic exposures, which retrospective biopsy collections cannot provide, and broadly reproduced the clinicopathologic relationships of measured spatial transcriptomics across 476 independent participants. Recent histology-based methods predict targeted gene panels at single-cell resolution,^47,48^ localize bulk RNA expression within healthy tissues,^49^ or predict expression across organs without retraining,^25,26^ and were developed predominantly on cancer or healthy tissue. In contrast, DeepSpot-M Kidney was adapted to diseased kidney tissue and remained more accurate than STPath and SQUALL, also after their fine-tuning on the same training biopsies. Moreover, most of these methods, like earlier predictors trained on single cohorts,^20,22,50^ predict a fixed set of genes represented during training, whereas DeepSpot-M queries each gene by an embedding of its protein sequence.^24^ This flexibility allowed the kidney-adapted model to extend Xenium panels to unmeasured genes and to profile rat kidneys without species-specific training or mapping to human orthologs, so that virtual spatial transcriptomics also applies to preclinical toxicology studies. Furthermore, accuracy was still increasing at 147 training biopsies, indicating that larger paired cohorts could further improve virtual spatial transcriptomics of the kidney.

The central finding is an axis of proximal tubular differentiation, along which virtual profiles reproduced lower gluconeogenic enzyme expression and higher injury and maladaptive repair markers in AKI and CKD than in DM-R. Across the atlas, higher proximal tubule program scores accompanied higher eGFR and less interstitial fibrosis and tubular atrophy. Together, these findings link the loss of differentiated proximal tubular expression, including gluconeogenic enzymes, to tubular injury, fibrosis, and lower kidney function.^4,10,51^ Studies of urinary EGF and renal gluconeogenesis described differentiated tubular expression as a marker of tubular health,^10,35^ and here this axis was read from routine histology. In parallel, injured tubule, fibrotic, and lymphoid aggregate niches expanded with interstitial fibrosis.

The main translational opportunity lies in existing biopsy archives, in which H&E slides are linked to eGFR trajectories, kidney failure, and death. Measured spatial transcriptomics is impractical at this scale, whereas virtual profiling needs only digitized archival H&E slides. Such archives would allow testing whether the molecular state of the tissue predicts outcomes beyond interstitial fibrosis, tubular atrophy, and eGFR,^35^ and searching for molecular subgroups within morphologically defined diagnoses, such as diabetic kidney disease.^38^ Moreover, virtual proximal tubule scores were associated with eGFR beyond interstitial fibrosis and tubular atrophy, suggesting that virtual profiles might capture a loss of differentiated tubular expression in tissue that still appears preserved, which matters when a needle biopsy samples relatively preserved cortex. Diagnostic applications, such as complementing chronicity scores or the automated Banff lesion scores now emerging in transplant pathology,^52^ remain speculative and will require prospective validation in several centers.

This study has several limitations, most notably the variable accuracy of virtual profiles across genes and programs. Immune and injury programs were reproduced least accurately, particularly genes that Visium rarely detected, such as *HAVCR1* (Figure 2b; Supplementary Table S7), so the inflammatory findings above are supportive rather than definitive. Deconvolution also indicated that virtual profiles underestimate adaptive tubular states in AKI (Supplementary Figure S6), and because the measured fractions are themselves estimates, we interpret virtual composition only in relative terms. Moreover, virtual profiles are derived from the same histology that pathologists grade, so part of their agreement with histopathologic descriptors is expected. In contrast, kidney function and, in rats, BUN and tissue expression provide independent evidence.

Each spot also combines tissue composition with cellular expression, so a lower program score cannot distinguish the loss of a cell population from altered expression in surviving cells. Single-cell virtual profiling may help separate the two.^23^ Furthermore, both the paired cohort and the atlas come from a single consortium that excludes glomerular diseases and transplants and relies on needle biopsies with little medulla, so the findings may not transfer to other institutions, diagnoses, or medullary programs. Rodent models reflect human kidney disease only partially,^53^ all rats were male, and small dose groups and arrays from separate kidney fragments restrict the rat results to validation under controlled exposure rather than prediction of human drug toxicity. In the atlas, FFPE tissue was validated against Xenium through niche shares and withheld panel genes rather than against Visium (Figure 4f and g), the associations are cross-sectional, and the gene-level eGFR associations remain exploratory. Therefore, cross-site molecular validation and longitudinal studies are needed before use beyond exploratory tissue characterization.

In conclusion, kidney-adapted virtual spatial transcriptomics opens existing, clinically annotated biopsy archives to spatial molecular analysis, complementing measured spatial transcriptomics in the study of disease-associated tissue programs. Linked to adjudicated diagnoses, histopathology, and kidney function, the virtual atlas provides a resource for generating and testing hypotheses on the tissue basis of kidney disease, selecting patients for targeted molecular profiling, and integrating spatial molecular readouts into translational studies.

## Disclosure

V.H.K. reports being an invited speaker for Sharing Progress in Cancer Care (SPCC) and Indica Labs; advisory board of Takeda; and sponsored research agreements with Roche and IAG, all unrelated to the current study. V.H.K. is a participant in a patent application on the assessment of cancer immunotherapy biomarkers by digital pathology; a patent application on multimodal deep learning for the prediction of recurrence risk in cancer patients, and a patent application on predicting the efficacy of cancer treatment using deep learning, all unrelated to the current work. G.R. is a participant in a patent application on matching cells from different measurement modalities which is not directly related to the current work. Moreover, G.R. is a cofounder of Computomics GmbH, Germany, and one of its shareholders. M.B.M. reports grants from the NovoNordisk Nordic Foundation, the Karolinska Institute Research Foundation, the Burgergemeinde Bern Foundation, and the Swiss National Science Foundation. All the other authors declared no competing interests.

## Data Sharing Statement

Previously published data were used for this study. Data Type: spatial transcriptomics, H&E whole-slide images, pathologist descriptors, and open-access clinical data of KPMP participants. Repository Name: KPMP Atlas Repository (https://atlas.kpmp.org; https://www.kpmp.org/available-data).^31^ Data Type: rat kidney H&E images, histopathology, clinical chemistry, and kidney expression arrays. Repository Name: Open TG-GATEs, Toxicogenomics Project and Toxicogenomics Informatics Project, CC BY-SA 2.1 JP (https://doi.org/10.18908/lsdba.nbdc00954-02-000; https://doi.org/10.18908/lsdba.nbdc00954-01-000).^30,39,40^ Original data generated for the study are available in a public access repository. Data Type: KPMP virtual spatial transcriptomics atlas (virtual expression of 19 338 genes per virtual spatial spot, tissue niche assignments, and slide metadata permitted for redistribution); virtual expression of the cells of the KPMP Xenium sections; per-gene accuracy in held-out KPMP biopsies; source data of all figures; code for the statistical analyses and figures. Repository Name: GitHub (https://github.com/ratschlab/projects2026-kidney-kpmp-analysis) and Hugging Face (https://huggingface.co/datasets/ratschlab/KPMP_virtual_spatial_transcriptomics_atlas; https://huggingface.co/datasets/ratschlab/KPMP_Xenium_virtual_spatial_transcriptomics).

## Data Availability

Previously published data were used for this study. Data Type: Spatial transcriptomics, hematoxylin and eosin (H&E) whole-slide images, pathologist descriptors, and open-access clinical data of KPMP participants. Repository Name: KPMP Atlas Repository (https://atlas.kpmp.org; https://www.kpmp.org/available-data). Data Type: Rat kidney H&E images, histopathology, clinical chemistry, and kidney expression arrays. Repository Name: Open TG-GATEs, Toxicogenomics Project and Toxicogenomics Informatics Project, CC BY-SA 2.1 JP (https://doi.org/10.18908/lsdba.nbdc00954-02-000; https://doi.org/10.18908/lsdba.nbdc00954-01-000). Original data generated for the study are available in a public access repository. Data Type: KPMP virtual spatial transcriptomics atlas (virtual expression of 19,338 genes per virtual spatial spot, tissue niche assignments, and slide metadata permitted for redistribution); virtual expression of the cells of the KPMP Xenium sections; per-gene accuracy in held-out KPMP biopsies; source data of all figures; code for the statistical analyses and figures. Repository Name: GitHub (https://github.com/ratschlab/projects2026-kidney-kpmp-analysis) and Hugging Face (https://huggingface.co/datasets/ratschlab/KPMP_virtual_spatial_transcriptomics_atlas; https://huggingface.co/datasets/ratschlab/KPMP_Xenium_virtual_spatial_transcriptomics).

https://doi.org/10.18908/lsdba.nbdc00954-02-000

https://doi.org/10.18908/lsdba.nbdc00954-01-000

https://huggingface.co/datasets/ratschlab/KPMP_virtual_spatial_transcriptomics_atlas

https://huggingface.co/datasets/ratschlab/KPMP_Xenium_virtual_spatial_transcriptomics

## Acknowledgments

We thank Hans-Peter Marti for his critical reading of the manuscript and helpful comments. We also thank the staff of the Swiss National Supercomputing Centre (CSCS) for their support on Alps (https://www.cscs.ch/computers/alps) within the Swiss AI Initiative (https://swiss-ai.org). Computational data analysis was performed at Leonhard Med (https://sis.id.ethz.ch/services/sensitiveresearchdata/), a secure trusted research environment at ETH Zurich, and we thank the Scientific IT Services of ETH Zurich for their support. This study uses data generated by the Kidney Precision Medicine Project (KPMP), and we acknowledge the support behind that data as follows: The results here are in whole or part based upon data generated by the KPMP, accessed August 3, 2026. https://www.kpmp.org. The Kidney Precision Medicine Project (KPMP) is supported by the National Institute of Diabetes and Digestive and Kidney Diseases (NIDDK) through the following grants: U01DK133081, U01DK133091, U01DK133092, U01DK133093, U01DK133095, U01DK133097, U01DK114866, U01DK114908, U01DK133090, U01DK133113, U01DK133766, U01DK133768, U01DK114907, U01DK114920, U01DK114923, U01DK114933, U24DK114886, UH3DK114926, UH3DK114861, UH3DK114915, UH3DK114937, 1U01DK144965-01, and 1U01DK144994-01. The present study received no funding from NIDDK, but we acknowledge the contributions of KPMP patient participants. Rat data are from Open TG-GATEs (Toxicogenomics Project and Toxicogenomics Informatics Project).

## Author Contributions

**Conceptualization:** Kalin Nonchev, Viktor H. Koelzer, Gunnar Rätsch, Matthias B. Moor. **Data curation:** Kalin Nonchev, Matthias B. Moor. **Formal analysis:** Kalin Nonchev. **Funding acquisition:** Viktor H. Koelzer, Gunnar Rätsch. **Investigation:** Kalin Nonchev, Yunsu Jang, Stellor Nlandu Khodo, Matthias B. Moor. **Methodology:** Kalin Nonchev. **Resources:** Kalin Nonchev, Viktor H. Koelzer, Gunnar Rätsch, Matthias B. Moor. **Software:** Kalin Nonchev. **Supervision:** Kalin Nonchev, Viktor H. Koelzer, Gunnar Rätsch, Matthias B. Moor. **Visualization:** Kalin Nonchev. **Writing – original draft:** Kalin Nonchev, Viktor H. Koelzer, Gunnar Rätsch, Matthias B. Moor. **Writing – review & editing:** Kalin Nonchev, Yunsu Jang, Stellor Nlandu Khodo, Viktor H. Koelzer, Gunnar Rätsch, Matthias B. Moor.

## Declaration of Generative AI and AI-Assisted Technologies in the Manuscript Preparation Process

During the preparation of this work the authors used Claude (Opus 5.5; Anthropic) and ChatGPT (5.5 Luna; OpenAI) in order to assist with prose editing, literature synthesis, and software development. After using these tools, the authors reviewed and edited the content as needed and take full responsibility for the content of the published article.

## References

[1] de Boer IH, Alpers CE, Azeloglu EU, et al. Rationale and design of the Kidney Precision Medicine Project. Kidney Int. 2021;99(3):498–510. doi:10.1016/j.kint.2020.08.039.

[2] Lake BB, Menon R, Winfree S, et al. An atlas of healthy and injured cell states and niches in the human kidney. Nature. 2023;619(7970):585–594. doi:10.1038/s41586-023-05769-3.

[3] Lake BB, Melo Ferreira R, Hansen J, et al. Cellular and Spatial Drivers of Unresolved Injury and Functional Decline in the Human Kidney. bioRxiv. 2025. doi:10.1101/2025.09.26.678707.

[4] Kirita Y, Wu H, Uchimura K, et al. Cell profiling of mouse acute kidney injury reveals conserved cellular responses to injury. Proceedings of the National Academy of Sciences. 2020;117(27):15874–15883. doi:10.1073/pnas.2005477117.

[5] Ferreira RM, Sabo AR, Winfree S, et al. Integration of spatial and single-cell transcriptomics localizes epithelial cell–immune cross-talk in kidney injury. JCI Insight. 2021;6(12). doi:10.1172/jci.insight.147703.

[6] Dixon EE, Wu H, Muto Y, et al. Spatially Resolved Transcriptomic Analysis of Acute Kidney Injury in a Female Murine Model. J Am Soc Nephrol. 2022;33(2):279–289. doi:10.1681/asn.2021081150.

[7] Isnard P, Li D, Xuanyuan Q, et al. Histopathologic Analysis of Human Kidney Spatial Transcriptomics Data. Am J Pathol. 2025;195(1):69–88. doi:10.1016/j.ajpath.2024.06.011.

[8] Abedini A, Levinsohn J, Klötzer KA, et al. Single-cell multi-omic and spatial profiling of human kidneys implicates the fibrotic microenvironment in kidney disease progression. Nat Genet. 2024;56(8):1712–1724. doi:10.1038/s41588-024-01802-x.

[9] Zhou D, Tian Y, Sun L, et al. Matrix Metalloproteinase-7 Is a Urinary Biomarker and Pathogenic Mediator of Kidney Fibrosis. J Am Soc Nephrol. 2017;28(2):598–611. doi:10.1681/asn.2016030354.

[10] Verissimo T, Faivre A, Rinaldi A, et al. Decreased Renal Gluconeogenesis Is a Hallmark of Chronic Kidney Disease. J Am Soc Nephrol. 2022;33(4):810–827. doi:10.1681/asn.2021050680.

[11] Huo Y, Deng R, Liu Q, et al. AI applications in renal pathology. Kidney Int. 2021;99(6):1309–20. doi:10.1016/j.kint.2021.01.015.

[12] Hermsen M, de Bel T, den Boer M, et al. Deep Learning–Based Histopathologic Assessment of Kidney Tissue. J Am Soc Nephrol. 2019;30(10):1968–1979. doi:10.1681/asn.2019020144.

[13] Bouteldja N, Klinkhammer BM, Bülow RD, et al. Deep Learning–Based Segmentation and Quantification in Experimental Kidney Histopathology. J Am Soc Nephrol. 2021;32(1):52–68. doi:10.1681/asn.2020050597.

[14] Ginley B, Jen KY, Han SS, et al. Automated Computational Detection of Interstitial Fibrosis, Tubular Atrophy, and Glomerulosclerosis. J Am Soc Nephrol. 2021 Feb;32(4):837–850. doi:10.1681/asn.2020050652.

[15] Barisoni L, Lafata KJ, Hewitt SM, et al. Digital pathology and computational image analysis in nephropathology. Nat Rev Nephrol. 2020;16(11):669–685. doi:10.1038/s41581-020-0321-6.

[16] Becker JU, Mayerich D, Padmanabhan M, et al. Artificial intelligence and machine learning in nephropathology. Kidney Int. 2020;98(1):65–75. doi:10.1016/j.kint.2020.02.027.

[17] Hölscher DL, Bouteldja N, Joodaki M, et al. Next-Generation Morphometry for pathomics-data mining in histopathology. Nat Commun. 2023;14(1). doi:10.1038/s41467-023-36173-0.

[18] Abe M, Niioka H, Matsumoto A, et al. Self-Supervised Learning for Feature Extraction from Glomerular Images and Disease Classification with Minimal Annotations. J Am Soc Nephrol. 2025 Oct;36(3):471–486. doi:10.1681/asn.0000000514.

[19] Li X, Shah M, Liu Q, et al. Computational Characterization of Lymphocyte Topology on Whole Slide Images of Glomerular Diseases. J Am Soc Nephrol. 2026 Oct;37(3):545–559. doi:10.1681/asn.0000000889.

[20] He B, Bergenstråhle L, Stenbeck L, et al. Integrating spatial gene expression and breast tumour morphology via deep learning. Nat Biomed Eng. 2020;4(8):827–34. doi:10.1038/s41551-020-0578-x.

[21] Schmauch B, Romagnoni A, Pronier E, et al. A deep learning model to predict RNA-Seq expression of tumours from whole slide images. Nat Commun. 2020;11(1):3877. doi:10.1038/s41467-020-17678-4.

[22] Nonchev K, Dawo S, Silina K, et al. DeepSpot: Leveraging spatial context for enhanced spatial transcriptomics prediction from H&E images. medRxiv. 2025. doi:10.1101/2025.02.09.25321567.

[23] Nonchev K, Manaiev G, Koelzer VH, Rätsch G. DeepSpot2Cell: Predicting virtual single-cell spatial transcriptomics from H&E images using spot-level supervision. bioRxiv. 2025. doi:10.1101/2025.09.23.678121.

[24] Nonchev K, Dawo S, Silina K, et al. DeepSpot-M: a multimodal foundation model for transcriptome-wide virtual spatial transcriptomics from histology. medRxiv. 2026. doi:10.64898/2026.06.19.26356060.

[25] Huang T, Liu T, Babadi M, et al. STPath: a generative foundation model for integrating spatial transcriptomics and whole-slide images. NPJ Digit Med. 2025;8(1):659. doi:10.1038/s41746-025-02020-3.

26. Zhang Z, Qin B, Zhao Y, et al. A multimodal foundation model to integrate histology with spatial molecular programs. bioRxiv. 2026. doi:10.64898/2026.06.01.729028.

[27] Shulman ED, Campagnolo EM, Lodha R, et al. AI-predicted spatial transcriptomics unlocks breast cancer biomarkers from pathology. Cell. 2026;189(14):4225–40.e25. doi:10.1016/j.cell.2026.04.023.

[28] Elnaggar A, Heinzinger M, Dallago C, et al. ProtTrans: toward understanding the language of life through self-supervised learning. IEEE Transactions on Pattern Analysis and Machine Intelligence. 2022;44(10):7112–27. doi:10.1109/TPAMI.2021.3095381.

[29] Hu D, Szklarczyk D, von Mering C, Jensen LJ. SPACE: STRING proteins as complementary embeddings. Bioinformatics. 2025;41(9):btaf496. doi:10.1093/bioinformatics/btaf496.

[30] Igarashi Y, Nakatsu N, Yamashita T, et al. Open TG-GATEs: a large-scale toxicogenomics database. Nucleic Acids Res. 2015;43(D1):D921–7. doi:10.1093/nar/gku955.

[31] [dataset] Kidney Precision Medicine Project. KPMP Kidney Tissue Atlas data repository; 2026. https://atlas.kpmp.org/repository. Accessed August 3, 2026.

[32] Karasikov M, van Doorn J, Känzig N, et al. Training state-of-the-art pathology foundation models with orders of magnitude less data. Medical Image Computing and Computer Assisted Intervention – MICCAI 2025, Lecture Notes in Computer Science. 2025:573–83. doi:10.1007/978-3-032-04984-1_55.

[33] You Y, Fu Y, Li L, et al. Systematic comparison of sequencing-based spatial transcriptomic methods. Nat Methods. 2024;21(9):1743–54. doi:10.1038/s41592-024-02325-3.

[34] Rademacher A, Huseynov A, Bortolomeazzi M, et al. Comparison of spatial transcriptomics technologies using tumor cryosections. Genome Biol. 2025;26(1):176. doi:10.1186/s13059-025-03624-4.

[35] Ju W, Nair V, Smith S, et al. Tissue transcriptome-driven identification of epidermal growth factor as a chronic kidney disease biomarker. Sci Transl Med. 2015;7(316). doi:10.1126/scitranslmed.aac7071.

[36] Schaub JA, Menon R, Melo Ferreira R, et al. Acute tubular injury versus acute interstitial nephritis in the kidney precision medicine project. JCI Insight. 2026;11(15). doi:10.1172/jci.insight.204127.

[37] Moledina DG, Obeid W, Smith RN, et al. Identification and validation of urinary CXCL9 as a biomarker for diagnosis of acute interstitial nephritis. J Clin Invest. 2023;133(13). doi:10.1172/jci168950.

[38] Dumoulin B, Levinsohn J, Klötzer KA, et al. Spatial atlas of diabetic kidney disease reveals a B cell-rich subgroup. Nature. 2026;653(8116):1158–1169. doi:10.1038/s41586-026-10363-4.

[39] [dataset] Toxicogenomics Project and Toxicogenomics Informatics Project. Open TG-GATEs; 2015. LSDB Archive. doi:10.18908/lsdba.nbdc00954-01-000.

[40] [dataset] Toxicogenomics Project and Toxicogenomics Informatics Project. Open TG-GATEs Pathological Image Database; 2015. LSDB Archive. doi:10.18908/lsdba.nbdc00954-02-000.

[41] Szklarczyk D, Kirsch R, Koutrouli M, et al. The STRING database in 2023: protein–protein association networks and functional enrichment analyses for any sequenced genome of interest. Nucleic Acids Res. 2023;51(D1):D638–46. doi:10.1093/nar/gkac1000.

[42] Irizarry RA, Hobbs B, Collin F, et al. Exploration, normalization, and summaries of high density oligonucleotide array probe level data. Biostatistics. 2003;4(2):249–64. doi:10.1093/biostatistics/4.2.249.

[43] Korsunsky I, Millard N, Fan J, et al. Fast, sensitive and accurate integration of single-cell data with Harmony. Nat Methods. 2019;16(12):1289–96. doi:10.1038/s41592-019-0619-0.

[44] Uehara T, Yamate J, Torii M, Maruyama T. Comparative nephrotoxicity of cisplatin and nedaplatin: mechanisms and histopathological characteristics. J Toxicol Pathol. 2011;24(2):87–94. doi:10.1293/tox.24.87.

[45] Murray G, Wyllie RG, Hill GS, et al. Experimental papillary necrosis of the kidney. I. Morphologic and functional data. Am J Pathol. 1972;67(2):285–302.

[46] Wedeen RP, Batuman V, Cheeks C, et al. Transport of gentamicin in rat proximal tubule. Lab Invest. 1983;48(2):212–23.

[47] Fu X, Cao Y, Bian B, et al. Spatial gene expression at single-cell resolution from histology using deep learning with GHIST. Nat Methods. 2025;22(9):1900–10. doi:10.1038/s41592-025-02795-z.

[48] Chadoutaud L, Lerousseau M, Herrero-Saboya D, et al. sCellST predicts single-cell gene expression from H&E images. Nat Commun. 2026;17:1194. doi:10.1038/s41467-025-67965-1.

[49] Cisternino F, Ometto S, Chatterjee S, et al. Self-supervised learning for characterising histomorphological diversity and spatial RNA expression prediction across 23 human tissue types. Nat Commun. 2024;15:5906. doi:10.1038/s41467-024-50317-w.

[50] Schmauch B, Herpin L, Olivier A, et al. A deep learning-based multiscale integration of spatial omics with tumor morphology. Nat Commun. 2025;16:11674. doi:10.1038/s41467-025-66691-y.

[51] Legouis D, Ricksten SE, Faivre A, et al. Altered proximal tubular cell glucose metabolism during acute kidney injury is associated with mortality. Nat Metab. 2020;2(8):732–743. doi:10.1038/s42255-020-0238-1.

[52] Buzzanca G, Pala C, He J, et al. BanffNET, a Deep Learning System for Comprehensive Histological Lesion Quantification in Kidney Transplant Biopsies. medRxiv. 2026. doi:10.64898/2026.08.28.26360029.

[53] Klötzer KA, Abedini A, Shen L, et al. Analysis of individual patient pathway coordination in a cross-species single-cell kidney atlas. Nat Genet. 2025;57(8):1922–34. doi:10.1038/s41588-025-02285-0.

