## supplement for "Virtual spatial transcriptomics links kidney histopathology to disease-associated molecular programs"

### Table of Contents

Supplementary Methods 2
Supplementary Table S1. Visium data of the paired KPMP specimens 7
Supplementary Table S2. Gene sets of nephron compartments and tissue programs 8
Supplementary Table S3. Accuracy against individual-spot and reference expression 9
Supplementary Table S4. Spearman correlation against individual-spot and reference expression 10
Supplementary Table S5. Accuracy on the most variable genes 11
Supplementary Table S6. Accuracy on the most spatially autocorrelated genes 12
Supplementary Table S7. Accuracy of the program genes 13
Supplementary Table S8. Virtual programs and eGFR beyond histopathology 14
Supplementary Table S9. Withheld Xenium panel genes by resolution, enrollment group, and Xenium panel 15
Supplementary Figure S1. Cohort, participant-level splits, and Visium data 16
Supplementary Figure S2. DeepSpot-M Kidney and the group-specific models 17
Supplementary Figure S3. Training curves and per-biopsy accuracy 18
Supplementary Figure S4. Accuracy as a function of the number of training biopsies 19
Supplementary Figure S5. Marker genes of program compartments in measured and virtual profiles 20
Supplementary Figure S6. Cell type fractions estimated by deconvolution 21
Supplementary Figure S7. Virtual tissue programs, kidney function, and adjudicated diagnoses 22
Supplementary Figure S8. Dose and time responses for five nephrotoxic compounds in rats 23
Supplementary Figure S9. Virtual versus measured kidney transcription in rats 24
Supplementary Figure S10. Example slides from the virtual atlas 25
Supplementary Figure S11. Scanner variation, niche composition, and gene-level eGFR associations in the virtual atlas 26
Supplementary Figure S12. Cell types and adaptive cell states measured by Xenium 27
Supplementary References 27

#### Supplementary Methods

##### Study population and data

The healthy reference tissue comprised 21 living-donor preperfusion biopsies, five nephrectomies, and two intraoperative biopsies, and all AKI, CKD, and DM-R specimens except one were percutaneous needle biopsies. The specimens comprised 179 226 spots (median, 850 per specimen; range, 78–4 060), and H&E images had 0.32 µm per pixel (168 specimens) or 0.64 µm per pixel (41 specimens; Supplementary Table S1 and Supplementary Figure S1). Clinical information came from the KPMP open-access participant table (accessed August 3, 2026), which recorded participant sex as sex at birth (female or male) and the race categories used to describe the cohort. Baseline eGFR was available as a continuous value and other clinical variables in categories. For participants with AKI, KPMP calculated baseline eGFR from the median of the last three outpatient creatinine values 7 to 365 days before enrollment, that is, before the injury episode.

##### DeepSpot-M and kidney adaptation

DeepSpot-M predicts the expression of a gene in an H&E tile from a query derived from a frozen embedding of the gene, here the ProtT5 embedding of the encoded protein,[S1,S2] which attends to the patch tokens of the pathology foundation model Midnight.[S3,S4] Gene-specific readout weights are generated from the same embedding, so any gene with an embedding can be queried. Adaptation started from the pretrained DeepSpot-M checkpoint and updated the decoder layers by minimizing the mean squared error between virtual and measured log-normalized expression of the 19 338 vocabulary genes. The decoder was trained with AdamW (learning rate $2 \times 10^{-4}$, weight decay $10^{-6}$, cosine decay to 1% of the initial rate) on batches of 35 tiles for a budget of 25 000 steps rounded to whole epochs (two epochs on the 147 training biopsies of each split and one epoch on all 209 biopsies), and the final checkpoint was used.

##### Accuracy metrics

Nearest neighbors for the reference expression were found in the space of the first 50 principal components of the measured log-normalized profiles of each biopsy and were shared by all models. Correlations were undefined, and therefore omitted, for genes without measured variance on a biopsy. Gene sets were summarized by the mean accuracy of their genes after averaging over groups and splits, and immune cell markers in the same way, namely macrophage (*AIF1*, *TYROBP*, *C1QA*, *C1QB*, *C1QC*, *CD14*, *LYZ*, *CD68*, *MRC1*, *CD163*, and *FCGR3A*), T cell (*CD3E*, *CD3D*, *CD2*, *CD4*, *CD8A*, *IL7R*, *CCL5*, *GZMK*, *NKG7*, and *GNLY*), and B cell markers (*MS4A1*, *CD79A*, *CD79B*, *CD19*, and *BANK1*). The mean accuracy of DeepSpot-M Kidney was 0.31 for macrophage, 0.18 for T cell, and 0.13 for B cell markers. The chance level of Figure 2a is the top-$K$ accuracy after permuting the virtual spots within each held-out biopsy (mean of 50 permutations, averaged over models). For gene rankings independent of held-out accuracy, the 15 542 genes predicted by every model were ranked on the training biopsies of each split in two ways. First, Seurat v3 highly variable gene analysis (scanpy) gave each gene's normalized variance within each biopsy, and genes were ordered by the number of biopsies in which they ranked among the 5 000 most variable, then by their median rank (four biopsies whose variance fit failed were left out; Supplementary Table S5). Second, genes were ordered by their mean Moran's $I$ of log-normalized expression on the six-nearest-spot graph of each biopsy (Supplementary Table S6).

##### Training-set size

The 147-biopsy training pool of split 0 was put in one group-stratified random order per subset seed (three seeds), in which each next biopsy came from the group furthest below its share of the pool, and the model with $n$ biopsies used the first $n$, so that subsets were nested and kept the group composition of the pool as far as $n$ allowed. Every model used the same step budget as the benchmark, so the model with all 147 biopsies corresponds to DeepSpot-M Kidney of split 0. Accuracy was expressed as the percentage gain over DeepSpot-M (zero-shot) in the same group, $(r_n/r_0 - 1) \times 100$, where $r_0$ is the zero-shot correlation, and the log-linear relation gain $= a + b \log_2 n$ was fitted by least squares to every subset order and $n \geq 1$, so that $b$ is the gain per doubling.

#### STPath and SQUALL

STPath[S5] and SQUALL[S6] were applied as released, without further training, to H&E patches centered on each spot, with the published code and checkpoints. STPath predicts spot expression from Prov-GigaPath tile features[S7] of all spots of a slide, here with kidney as organ and Visium as technology, and SQUALL predicts expression on fine bins, which we averaged over each spot. Each model was scored against the same measured targets as DeepSpot-M on the genes it shared with DeepSpot-M. Of the 21 KPMP specimens in the published training collections of both models,[S8] the 19 held out at least once in our splits were excluded from the top-$K$ comparison for all models (Figure 2a and Supplementary Tables S3 and S4).

#### Fine-tuned STPath and SQUALL

STPath and SQUALL were also fine-tuned on the 147 training biopsies of each split, starting from the released weights and following each model's published training objective and optimizer, namely masked prediction of spot expression from Prov-GigaPath patch features for STPath[S5] and reconstruction of masked image tokens and binarized expression maps for SQUALL.[S6] For both models, the training length was selected on a validation tenth of the training biopsies, with SQUALL limited to the tile budget of DeepSpot-M Kidney, and the model was then retrained on all training biopsies. Virtual expression of the held-out biopsies was scored as for the released models (Figure 2a).

#### Program compartments and cell type fractions

*Program compartments.* Measured and virtual expression of each spot of the 171 held-out biopsies was scored for the eight atlas programs of Supplementary Table S2 as the mean log1p expression of program genes minus that of expression-matched control genes, as in the atlas. Program scores were $z$-scored within each source, and each spot was assigned to the program with the highest score. Canonical marker genes of kidney cell types, supplemented from the KPMP single-cell atlas,[S8] were summarized per compartment and disease group by their mean $z$-score and the percentage of spots above the gene's 75th percentile. Specificity was assessed with the markers outside the program gene sets, by the area under the receiver operating characteristic curve (AUROC) of their mean $z$-score for spots of a compartment versus all other spots, with 95% confidence intervals from bootstrap resampling of biopsies, and by the number of markers whose highest mean $z$-score fell in their own compartment. Virtual marker scores were also evaluated on the compartments assigned from measured expression.

*Cell type fractions.* Spots were deconvolved with the KPMP single-cell RNA sequencing atlas[S8] (CELLxGENE release 1.0, 84 818 cells in 14 classes from all enrollment groups after removal of degenerative and cycling cells). The signature of each class was the mean expression (counts per 10 000) of its 50 top markers, ranked by the log2 ratio of their mean expression in the class to their highest mean expression in any other class (600 genes in total). Each spot was fitted by non-negative least squares[S9] on genes scaled to their maximum, using linear measured expression and back-transformed virtual expression, and the RNA shares were divided by the mean number of unique molecular identifiers per cell of each class[S10] and renormalized to cell fractions. Fractions were averaged per participant and then per group. On pseudo-spots of 4 to 10 cells simulated from held-out donors of the reference, the median per-class correlation between true and estimated cell fractions was 0.77, and the single-nucleus atlas (133 808 nuclei) served as a sensitivity reference.

#### Rat toxicology study

*Data.* Open TG-GATEs (Toxicogenomics Project-Genomics Assisted Toxicity Evaluation System)[S11] (pathological image database and clinical data; Toxicogenomics Project and Toxicogenomics Informatics Project, CC BY-SA 2.1 JP) provides kidney H&E sections of male Crj:CD(SD)IGS rats aged 6 weeks, scanned with an Aperio scanner at 20× (0.492–0.496 µm per pixel). Doses (low, middle, high; mg/kg) were 0.3, 1, and 3 (single) and 0.1, 0.3, and 1 (daily) for cisplatin; 10, 30, and 100 and 1, 3, and 10 for carboplatin; 10, 30, and 100 for gentamicin; 6, 20, and 60 and 2, 6, and 20 for 2-bromoethylamine; 12, 40, and 120 and 4, 12, and 40 for puromycin aminonucleoside; and 1, 3, and 10 and 0.1, 0.3, and 1 for doxorubicin. We included every animal with a kidney expression array (567 rats; 423 treated rats in 142 dose groups of up to three animals and 144 controls; fewer than three where animals died) and used one capture per animal showing the whole transverse section with papilla and pelvis. Histopathologic findings were graded minimal (1), slight (2), moderate (3), or severe (4). Findings flagged as spontaneous in the Open TG-GATEs pathology table were excluded, and 76 rats had a treatment-related finding, 35 of them moderate or severe.

*Virtual profiling.* DeepSpot-M Kidney adapted on all 209 paired biopsies was queried with the STRING v12 ProtT5 embeddings of 22 237 rat proteins (20 368 genes).[S1,S2,S12] Tiles with glass, ink, folds, or blur were removed (11.2 million tiles retained; median, 19 051 per rat), and the 65 µm crops (131–132 pixels) were resized to 224 pixels. Virtual log expression of each program gene was averaged per zone. Depth was computed on the largest tissue piece as $d_C/(d_C + d_P)$, with $d_C$ and $d_P$ the distances to the capsule and to the pelvic surface, and the cortex, outer medulla, and inner medulla with papilla were delimited at depths 0.49 and 0.76. These borders were the median depths of the steepest decline in eosin optical density (color deconvolution of the H&E thumbnail, per-section $z$-scores, 40 depth bins) in the 144 control sections, where eosinophilic cortex gives way to the outer medulla and the pale inner medulla begins. They agreed with the ends of the virtual glomerulus and thick ascending limb programs in the same sections (depths 0.46 and 0.76), and the cortical border ran through the arcuate vessels. Scanner stripes with a period of about 1 mm were removed from maps by subtracting a moving average of lattice columns.

*Readouts and statistics.* Time-matched controls had the same compound, regimen, and time, each control was compared with the other controls of its group, and the pooled within-group SD came from all 144 controls. Expected zones were taken from the literature before the analysis, namely outer medullary injury for cisplatin, extending to the cortex at higher doses;[S13,S14] papillary and inner medullary injury for 2-bromoethylamine, with outer medullary involvement;[S15,S16] and cortical proximal tubule loss for gentamicin, which accumulates in the S1 and S2 segments.[S17] Puromycin aminonucleoside and doxorubicin injure podocytes first, so cortical proximal tubule loss was expected only as a later, secondary change.[S18,S19] Carboplatin, which causes little kidney injury in rats, served as a low-nephrotoxicity comparator.[S20] Doxorubicin, with two rats with lesions, was not included in zone tests. Zone specificity in rats with lesions was tested by Wilcoxon signed-rank tests of the expected zone minus the mean of the other two. AUCs for histopathology (moderate or severe versus no finding, 35 versus 491) and for BUN at least twice the mean of the matched controls (13 versus 554), used the whole-section injury score, with 95% CIs from 2 000 bootstrap resamples drawn separately from the rats with and without the outcome.

*Kidney expression arrays.* The GeneChip Rat Genome 230 2.0 arrays (Affymetrix) of the 567 rats (left kidney, 1 mm fragments containing cortex and medulla) were normalized together by robust multi-array average.[S21] Probesets with a single gene symbol were kept, and the probeset with the highest mean expression represented each gene (14 603 genes). Measured and virtual whole-section scores used the program genes present in both, namely *Lcn2*, *Clu*, *Spp1*, *Timp1*, and *Cd44* for injury (*Sox9* has no probeset) and *Lrp2*, *Slc34a1*, and *Slc5a2* for proximal tubule loss, were standardized against the matched controls in the same way, and were compared by Spearman correlation, with 95% confidence intervals from 10 000 bootstrap resamples of the 48 control blocks. Partial correlations adjusted both ranks for the maximum treatment-related grade.

#### KPMP virtual spatial transcriptomics atlas

*Slides and prediction.* All H&E whole-slide images of the KPMP open-access repository (downloaded from October 23, 2026) of participants outside the Visium cohort were included. KPMP slides carry two to four serial levels of the core. Slides were scanned on two Leica Biosystems scanners, Aperio ScanScope (0.253 µm per pixel) and Aperio GT450 (0.262–0.263 µm per pixel), except for one FFPE slide. After tissue detection on a coarse pyramid level and removal of glass, ink, folds, blurred, and unstained tiles (up to 50 000 tiles per slide), the 65 µm crops (247–257 pixels) were resized to 224 pixels, yielding 5 458 825 virtual spots and 17 119 $mm^2$ of tissue.

*Program scores and associations.* Each program gene was compared with the mean virtual log1p expression of 100 random expression-matched control genes (24 expression bins). Pathologist descriptors were parsed as continuous percentages or ordinal grades and kept when available for at least 30 participants. Rank partial correlations residualized both ranked variables on site, sex, age bin, and scanner share (the fraction of a participant's slides scanned on the GT450), with cortex fraction added for descriptors, and FDR was controlled over the 72 cells shown per tissue preparation. Visium partial correlations used the same program scores, computed from measured pseudo-bulk profiles, and the same covariates except scanner. Associations of genes with eGFR used the 7 091 genes with mean virtual log1p expression of at least 0.1. The contribution of program scores to eGFR beyond histopathology (Supplementary Table S8) was assessed in the FFPE participants without AKI with interstitial fibrosis and tubular atrophy scores by repeated cross-validation (10 folds, 20 repeats) of ridge regression with the covariates, interstitial fibrosis, and tubular atrophy, with and without the 18 virtual program scores. $R^2$ was computed on the out-of-fold predictions of all participants in each repeat and averaged over the 20 repeats, and the standardization and the ridge penalty (25 values, efficient leave-one-out) were fitted within each training fold.

*Niches.* On GT450 FFPE slides, features were the atlas $z$-scores of 403 genes (the 103 program genes and the 300 nonribosomal, nonmitochondrial genes with the largest SD of virtual expression across the atlas), from which each spot's overall virtual expression level, the mean virtual log expression over all genes standardized within the scanner model, was regressed out with one least-squares slope per gene. Up to 300 spots per slide (174 900 spots) were reduced to 30 principal components, integrated across the four GT450 instruments with Harmony (default settings),[S22] clustered with the Leiden algorithm on a 15-nearest-neighbor graph (resolution 0.4), and embedded by UMAP, and every GT450 spot was assigned by a 15-nearest-neighbor vote among the clustered spots of the same instrument in the principal components before integration (agreement with the clustering, 0.97). A gene marked a niche when its mean exceeded that of every other niche by at least 0.5 SD. Representative tiles were drawn, one per participant, from the decile of non-edge, densely stained tiles closest to the niche centroid whose 15 nearest neighbors all belonged to the niche.

*Xenium validation.* KPMP Xenium sections of 49 participants (1.65 million cells; 24 sections on the hKidney and 25 on the hMulti 480-gene panel) were typed by marker scoring, the mean capped, panel-scaled expression of each cell type's markers, with the type of the highest score assigned and cells with fewer than 10 transcripts or a best score below 0.25 left unassigned. On the coarse classes of proximal tubule (including injured proximal tubule), thick ascending limb, and other cells, the marker calls agreed with a nearest-centroid transfer from the KPMP single-nucleus reference[S8] for 97.5% of the typed cells. Each section was divided into the 56 µm spot lattice of the atlas, each spot was assigned the atlas niche of its virtual expression, predicted from the H&E image of the section, by projection onto the atlas principal components centered per imaging center and the majority niche of its 15 nearest atlas spots, and the dominant cell type of a spot was the most frequent measured type among its typed cells. Niche shares of the atlas slides were compared with measured cell shares in the participants with both a Xenium section and GT450 FFPE slides. Adaptive cell states were assessed per enrollment group on the sections with at least 200 PT cells, as the share of injured PT cells among PT cells by marker scoring (*HAVCR1*, *VCAM1*, *CDH6*, and *DCC*) and as the shares of adaptive PT and adaptive TAL cells by nearest-centroid transfer from the KPMP single-nucleus reference,[S8] compared across groups by Kruskal–Wallis tests. The share of spots of each section in the virtual injured tubule niche was compared with the measured injured PT share by Spearman correlation.

*Virtual extension of Xenium panels.* The H&E image of each Xenium section was registered to the Xenium coordinates by an affine transformation, accepted when the median distance of Xenium nuclei to the nearest hematoxylin-stained nucleus was below 8 µm. Each cell with at least one transcript was represented by the 38 µm H&E crop centered on its centroid,[S4] and up to 50 000 cells per section, balanced across Leiden clusters of their measured expression, entered the adaptation. Of the 742 genes of the two panels in the model vocabulary, the 233 genes on chromosomes 1, 3, 5, 7, and 9 were held out for scoring, the 9 genes on other chromosomes with an Ensembl 113 within-species paralog of at least 25% identity among the held-out genes were withheld from adaptation and scoring,[S4] and the remaining genes of each section's panel entered the loss. Starting from DeepSpot-M Kidney adapted on all 209 Visium participants, only the expression decoder was trained on all 49 sections together with the optimizer, batch size, step budget, and final checkpoint of the Visium adaptation (one epoch). Virtual expression before and after adaptation was scored per section and held-out gene by the Pearson correlation across the cells of the section with the reference expression of the five nearest neighbors in expression space, as for Visium, and summarized by the median over genes on the 34 sections of participants outside the Visium training. The adaptation was repeated at the resolution of the atlas with the same held-out and withheld genes. For this, each section was divided into the 56 µm spot lattice of the atlas, the counts of the cells whose centroid fell into a lattice square were summed, and squares with at least three cells were kept, each with the 65 µm H&E field of view of the atlas centered on it, while training and scoring were unchanged. Both analyses are summarized against the expression of individual cells or patches, by enrollment group, and by Xenium panel in Supplementary Table S9.

#### Software

Analyses used Python 3.12.13 with PyTorch 2.8.0, PyTorch Lightning 2.6.1, timm 1.0.26, Scanpy 1.12.1,[S23] AnnData 0.12.10, leidenalg 0.11.0, harmonypy 2.1.0, UMAP 0.5.12, scikit-learn 1.8.0, SciPy 1.17.1, statsmodels 0.14.6, NumPy 2.4.4, pandas 2.3.3, OpenSlide 4.0.0, pyvips 2.2.3 (libvips 8.15.3), and WebDataset 0.2.100. Rat expression arrays were normalized in R 4.2.3 with affy 1.76.0.

**Supplementary Table S1. Visium data of the paired KPMP specimens.** The 209 frozen specimens with paired H&E images and 10x Genomics Visium profiles, one per participant, by enrollment group. Spots per specimen, median (range); UMIs and genes per spot, median over the spots of each specimen, then median (interquartile range) over specimens; genes detected, genes with at least one count; pixel size, specimens with 0.32 or 0.64 µm per pixel. Distributions in Supplementary Figure S1d–g. DM-R, diabetes mellitus resilient; UMI, unique molecular identifier.

| Group | Specimens | Spots | Spots per specimen | UMIs per spot | Genes per spot | Genes detected | Pixel size, 0.32 / 0.64 µm |
|---|---|---|---|---|---|---|---|
| AKI | 62 | 46 152 | 748 (294–1 399) | 7 677 (3 733–11 160) | 3 394 (2 111–4 181) | 20 608 (19 550–21 489) | 50 / 12 |
| CKD | 95 | 81 900 | 880 (132–2 053) | 5 326 (2 993–7 723) | 2 509 (1 649–3 212) | 20 320 (19 122–21 165) | 75 / 20 |
| DM-R | 24 | 21 155 | 910 (200–1 455) | 5 162 (3 486–7 016) | 2 406 (1 918–2 878) | 20 286 (19 324–21 036) | 23 / 1 |
| Reference | 28 | 30 019 | 884 (78–4 060) | 4 100 (2 803–7 162) | 2 288 (1 698–3 076) | 19 976 (18 228–20 904) | 20 / 8 |
| All | 209 | 179 226 | 850 (78–4 060) | 5 841 (3 216–8 588) | 2 633 (1 809–3 412) | 20 346 (19 131–21 189) | 168 / 41 |

**Supplementary Table S2. Gene sets of nephron compartments and tissue programs.** Gene sets of Figure 2b and Supplementary Figure S7, from published kidney atlas and Kidney Precision Medicine Project studies, and two sets used only in the atlas niche features and the program scores of Supplementary Table S8, adaptive PT[S8] and inflammatory fibroblast.[S24,S25] The atlas programs of Figure 4d use the compartment sets, except proximal tubule (*LRP2*, *CUBN*, *SLC34A1*, *ALDOB*, *GATM*, *GSTA1*, *SLC5A2*, *ACSM2A*, and *PLG*), injury and repair (the injury, repair PT score), and immune (the immune score). Held-out accuracy of the atlas programs (mean Pearson $r$ of DeepSpot-M Kidney against reference and, in parentheses, individual-spot expression): proximal tubule, 0.54 (0.28); loop of Henle and TAL, 0.46 (0.28); distal tubule and collecting duct, 0.44 (0.26); podocyte, 0.43 (0.21); vessel and smooth muscle, 0.55 (0.31); fibroblast, 0.38 (0.19); immune, 0.28 (0.13); injury and repair, 0.19 (0.07). IFN, interferon; PT, proximal tubule; TAL, thick ascending limb; TLS, tertiary lymphoid structure.

| Gene set | $n$ | Genes |
|---|---|---|
| *Compartments and programs (Figure 2B)* | | |
| Proximal tubule | 14 | *LRP2*, *CUBN*, *SLC34A1*, *ALDOB*, *GATM*, *GSTA1*, *SLC5A2*, *ACSM2A*, *PLG*, *THY1*, *MT1G*, *GPX3*, *APOE*, *PDZK1IP1* |
| PT gluconeogenesis | 4 | *PC*, *PCK1*, *FBP1*, *G6PC1* |
| Podocyte | 4 | *NPHS1*, *NPHS2*, *PODXL*, *WT1* |
| Loop of Henle, TAL | 3 | *UMOD*, *SLC12A1*, *EGF* |
| Distal tubule, collecting duct | 3 | *CALB1*, *AQP2*, *DEFB1* |
| Vessel, smooth muscle | 7 | *ACTA2*, *TAGLN*, *MYH11*, *AQP1*, *A2M*, *PDGFRB*, *VIM* |
| Fibroblast | 10 | *DCN*, *LUM*, *C7*, *PDGFRA*, *FBLN1*, *COL1A1*, *COL1A2*, *COL3A1*, *POSTN*, *FAP* |
| Myofibroblast | 3 | *INHBA*, *SPARC*, *TNC* |
| Injury, repair | 19 | *VCAM1*, *HAVCR1*, *MMP7*, *CDH6*, *CDKN1A*, *LCN2*, *SPP1*, *IGFBP7*, *SOX9*, *PROM1*, *DCDC2*, *ITGB8*, *ITGB6*, *CD24*, *TPM1*, *CLU*, *CST3*, *TIMP1*, *FN1* |
| Myeloid, IFN-$\gamma$, T cell | 14 | *PTPRC*, *CD68*, *MRC1*, *CCL2*, *C3*, *B2M*, *CXCL9*, *CXCL10*, *IFNG*, *TNF*, *CD3E*, *CD8A*, *STAT1*, *HLA-DRA* |
| B cell, TLS | 9 | *CD79A*, *MS4A1*, *CD19*, *MZB1*, *CXCL13*, *LTB*, *CCL19*, *CCL21*, *JCHAIN* |
| *Program scores (Supplementary Figure S7)* | | |
| Injury, repair PT | 5 | *VCAM1*, *HAVCR1*, *MMP7*, *CDH6*, *CDKN1A* |
| Adaptive TAL | 5 | *TMPRSS4*, *CREB5*, *ITGA3*, *ITGB6*, *ITGB8* |
| Healthy PT | 10 | *THY1*, *MT1G*, *GPX3*, *APOE*, *PDZK1IP1*, *GATM*, *ALDOB*, *LRP2*, *CUBN*, *SLC34A1* |
| PT gluconeogenesis | 4 | *PC*, *PCK1*, *FBP1*, *G6PC1* |
| EGF, UMOD | 2 | *EGF*, *UMOD* |
| Fibroblast content | 8 | *DCN*, *LUM*, *C7*, *PDGFRA*, *FBLN1*, *COL1A1*, *COL1A2*, *COL3A1* |
| Myofibroblast | 5 | *INHBA*, *SPARC*, *TNC*, *POSTN*, *FAP* |
| Immune | 6 | *PTPRC*, *CD68*, *MRC1*, *CCL2*, *C3*, *B2M* |
| IFN-$\gamma$, CXCL9 | 8 | *CXCL9*, *CXCL10*, *IFNG*, *TNF*, *CD3E*, *CD8A*, *STAT1*, *HLA-DRA* |
| B cell, TLS | 9 | *CD79A*, *MS4A1*, *CD19*, *MZB1*, *CXCL13*, *LTB*, *CCL19*, *CCL21*, *JCHAIN* |
| Degenerative | 5 | *GDF15*, *CDKN2A*, *NAMPT*, *CST3*, *CLU* |
| *Atlas only (niche features; Supplementary Table S8)* | | |
| Adaptive PT | 1 | *ROBO2* |
| Inflammatory fibroblast | 6 | *OSMR*, *IL1R1*, *SOX4*, *MID1*, *PXDN*, *CHI3L1* |

**Supplementary Table S3. Accuracy against individual-spot and reference expression.** Mean Pearson correlation of the *K* best-predicted genes, ranked independently for each model, on held-out biopsies per group outside the training data of SQUALL and STPath (mean over five splits), against individual-spot expression and, in parentheses, against reference expression (Methods; Figure 2a). Models as in Figure 2a, as released (DeepSpot-M without KPMP data, SQUALL and STPath with KPMP specimens in pretraining) or fine-tuned on the training biopsies of each split. DeepSpot-M fine-tuned (DeepSpot-M Kidney) in bold. DM-R, diabetes mellitus resilient; KPMP, Kidney Precision Medicine Project.

| Genes (*K*) | Model | AKI | CKD | DM-R | Reference |
|---|---|---|---|---|---|
| 100 | DeepSpot-M as released | 0.13 (0.28) | 0.15 (0.34) | 0.17 (0.41) | 0.14 (0.30) |
| | SQUALL as released | 0.07 (0.16) | 0.07 (0.17) | 0.10 (0.24) | 0.10 (0.21) |
| | STPath as released | 0.28 (0.53) | 0.27 (0.50) | 0.32 (0.63) | 0.28 (0.51) |
| | SQUALL fine-tuned | 0.09 (0.21) | 0.11 (0.26) | 0.13 (0.30) | 0.12 (0.25) |
| | STPath fine-tuned | 0.29 (0.55) | 0.29 (0.54) | 0.36 (0.66) | 0.29 (0.53) |
| | DeepSpot-M fine-tuned | **0.31 (0.56)** | **0.34 (0.60)** | **0.37 (0.65)** | **0.31 (0.55)** |
| 1 000 | DeepSpot-M as released | 0.07 (0.18) | 0.09 (0.22) | 0.10 (0.28) | 0.08 (0.19) |
| | SQUALL as released | 0.03 (0.08) | 0.04 (0.10) | 0.06 (0.14) | 0.06 (0.13) |
| | STPath as released | 0.15 (0.32) | 0.16 (0.34) | 0.18 (0.42) | 0.15 (0.33) |
| | SQUALL fine-tuned | 0.06 (0.15) | 0.08 (0.19) | 0.09 (0.22) | 0.08 (0.17) |
| | STPath fine-tuned | 0.16 (0.35) | 0.17 (0.37) | 0.20 (0.45) | 0.16 (0.35) |
| | DeepSpot-M fine-tuned | **0.19 (0.41)** | **0.20 (0.44)** | **0.22 (0.49)** | **0.18 (0.39)** |
| 5 000 | DeepSpot-M as released | 0.04 (0.11) | 0.05 (0.14) | 0.06 (0.17) | 0.05 (0.11) |
| | SQUALL as released | 0.02 (0.04) | 0.02 (0.05) | 0.03 (0.08) | 0.03 (0.07) |
| | STPath as released | 0.08 (0.18) | 0.09 (0.21) | 0.10 (0.25) | 0.08 (0.18) |
| | SQUALL fine-tuned | 0.04 (0.10) | 0.05 (0.13) | 0.06 (0.15) | 0.05 (0.11) |
| | STPath fine-tuned | 0.09 (0.21) | 0.09 (0.23) | 0.11 (0.27) | 0.08 (0.20) |
| | DeepSpot-M fine-tuned | **0.10 (0.25)** | **0.11 (0.28)** | **0.12 (0.30)** | **0.10 (0.24)** |
| All | DeepSpot-M as released | 0.02 (0.04) | 0.02 (0.06) | 0.02 (0.07) | 0.02 (0.03) |
| | SQUALL as released | 0.00 (0.00) | 0.00 (0.01) | 0.01 (0.02) | 0.00 (0.01) |
| | STPath as released | 0.03 (0.08) | 0.04 (0.09) | 0.04 (0.11) | 0.03 (0.07) |
| | SQUALL fine-tuned | 0.02 (0.04) | 0.02 (0.06) | 0.03 (0.07) | 0.02 (0.05) |
| | STPath fine-tuned | 0.04 (0.10) | 0.04 (0.11) | 0.05 (0.12) | 0.03 (0.09) |
| | DeepSpot-M fine-tuned | **0.05 (0.11)** | **0.05 (0.13)** | **0.05 (0.14)** | **0.04 (0.11)** |

**Supplementary Table S4. Spearman correlation against individual-spot and reference expression.** Mean Spearman correlation of the $K$ best-predicted genes, ranked independently for each model, on the held-out biopsies of Supplementary Table S3 (mean over five splits), against individual-spot expression and, in parentheses, against reference expression (Methods; Figure 2a). Models and bold as in Supplementary Table S3. DM-R, diabetes mellitus resilient.

| Genes ($K$) | Model | AKI | CKD | DM-R | Reference |
|---|---|---|---|---|---|
| 100 | DeepSpot-M as released | 0.13 (0.27) | 0.14 (0.31) | 0.17 (0.40) | 0.13 (0.26) |
| | SQUALL as released | 0.07 (0.15) | 0.08 (0.17) | 0.10 (0.22) | 0.10 (0.20) |
| | STPath as released | 0.20 (0.39) | 0.23 (0.42) | 0.25 (0.50) | 0.20 (0.37) |
| | SQUALL fine-tuned | 0.09 (0.19) | 0.10 (0.23) | 0.12 (0.26) | 0.12 (0.23) |
| | STPath fine-tuned | 0.22 (0.42) | 0.24 (0.45) | 0.27 (0.51) | 0.22 (0.39) |
| | DeepSpot-M fine-tuned | **0.30 (0.53)** | **0.33 (0.57)** | **0.34 (0.58)** | **0.27 (0.45)** |
| 1 000 | DeepSpot-M as released | 0.07 (0.16) | 0.09 (0.20) | 0.11 (0.26) | 0.08 (0.17) |
| | SQUALL as released | 0.04 (0.08) | 0.05 (0.10) | 0.06 (0.14) | 0.06 (0.12) |
| | STPath as released | 0.12 (0.26) | 0.14 (0.29) | 0.16 (0.34) | 0.12 (0.25) |
| | SQUALL fine-tuned | 0.06 (0.13) | 0.08 (0.17) | 0.09 (0.19) | 0.08 (0.16) |
| | STPath fine-tuned | 0.14 (0.29) | 0.16 (0.31) | 0.18 (0.36) | 0.13 (0.27) |
| | DeepSpot-M fine-tuned | **0.18 (0.36)** | **0.20 (0.40)** | **0.20 (0.41)** | **0.16 (0.32)** |
| 5 000 | DeepSpot-M as released | 0.04 (0.10) | 0.06 (0.13) | 0.07 (0.16) | 0.05 (0.10) |
| | SQUALL as released | 0.02 (0.04) | 0.03 (0.06) | 0.03 (0.07) | 0.03 (0.07) |
| | STPath as released | 0.08 (0.16) | 0.10 (0.19) | 0.11 (0.22) | 0.08 (0.15) |
| | SQUALL fine-tuned | 0.04 (0.09) | 0.06 (0.12) | 0.07 (0.14) | 0.06 (0.10) |
| | STPath fine-tuned | 0.09 (0.18) | 0.11 (0.21) | 0.12 (0.24) | 0.09 (0.17) |
| | DeepSpot-M fine-tuned | **0.11 (0.22)** | **0.13 (0.25)** | **0.13 (0.27)** | **0.10 (0.20)** |
| All | DeepSpot-M as released | 0.02 (0.04) | 0.03 (0.06) | 0.03 (0.07) | 0.02 (0.03) |
| | SQUALL as released | 0.00 (0.00) | 0.01 (0.01) | 0.01 (0.01) | 0.00 (0.01) |
| | STPath as released | 0.04 (0.07) | 0.05 (0.09) | 0.05 (0.11) | 0.03 (0.06) |
| | SQUALL fine-tuned | 0.02 (0.04) | 0.03 (0.07) | 0.03 (0.07) | 0.03 (0.04) |
| | STPath fine-tuned | 0.05 (0.09) | 0.06 (0.11) | 0.06 (0.12) | 0.04 (0.08) |
| | DeepSpot-M fine-tuned | **0.05 (0.11)** | **0.07 (0.13)** | **0.06 (0.14)** | **0.05 (0.10)** |

**Supplementary Table S5. Accuracy on the most variable genes.** Mean Pearson correlation of the $K$ genes with the highest Seurat v3 variability, with each training biopsy of a split as a batch, among the 15 542 genes predicted by every model, on the held-out biopsies of Supplementary Table S3 (mean over five splits), against individual-spot expression and, in parentheses, against reference expression (Supplementary Methods). Models and bold as in Supplementary Table S3. DM-R, diabetes mellitus resilient.

| Genes ($K$) | Model | AKI | CKD | DM-R | Reference |
|---|---|---|---|---|---|
| 100 | DeepSpot-M as released | 0.03 (0.08) | 0.04 (0.11) | 0.05 (0.13) | 0.03 (0.08) |
| | SQUALL as released | 0.00 (0.00) | 0.01 (0.01) | 0.02 (0.03) | 0.01 (0.01) |
| | STPath as released | 0.20 (0.37) | 0.20 (0.38) | 0.23 (0.45) | 0.18 (0.33) |
| | SQUALL fine-tuned | 0.03 (0.06) | 0.03 (0.08) | 0.04 (0.08) | 0.02 (0.05) |
| | STPath fine-tuned | 0.22 (0.41) | 0.22 (0.43) | 0.26 (0.49) | 0.19 (0.36) |
| | DeepSpot-M fine-tuned | **0.19 (0.38)** | **0.20 (0.41)** | **0.23 (0.47)** | **0.17 (0.35)** |
| 1 000 | DeepSpot-M as released | 0.03 (0.07) | 0.03 (0.09) | 0.04 (0.10) | 0.02 (0.05) |
| | SQUALL as released | 0.00 (0.00) | 0.00 (0.01) | 0.01 (0.02) | 0.00 (0.01) |
| | STPath as released | 0.11 (0.24) | 0.12 (0.26) | 0.12 (0.30) | 0.10 (0.22) |
| | SQUALL fine-tuned | 0.02 (0.05) | 0.03 (0.07) | 0.03 (0.07) | 0.02 (0.05) |
| | STPath fine-tuned | 0.13 (0.29) | 0.13 (0.30) | 0.15 (0.34) | 0.11 (0.25) |
| | DeepSpot-M fine-tuned | **0.13 (0.30)** | **0.14 (0.32)** | **0.15 (0.35)** | **0.12 (0.26)** |
| 5 000 | DeepSpot-M as released | 0.02 (0.06) | 0.03 (0.08) | 0.03 (0.08) | 0.02 (0.04) |
| | SQUALL as released | 0.00 (0.00) | 0.00 (0.00) | 0.00 (0.01) | 0.00 (0.01) |
| | STPath as released | 0.05 (0.13) | 0.06 (0.15) | 0.06 (0.17) | 0.05 (0.12) |
| | SQUALL fine-tuned | 0.02 (0.05) | 0.03 (0.07) | 0.03 (0.08) | 0.02 (0.05) |
| | STPath fine-tuned | 0.06 (0.16) | 0.07 (0.18) | 0.08 (0.20) | 0.06 (0.14) |
| | DeepSpot-M fine-tuned | **0.07 (0.18)** | **0.08 (0.20)** | **0.08 (0.21)** | **0.06 (0.16)** |
| All | DeepSpot-M as released | 0.02 (0.04) | 0.02 (0.06) | 0.02 (0.07) | 0.02 (0.03) |
| | SQUALL as released | 0.00 (0.00) | 0.00 (0.01) | 0.01 (0.02) | 0.00 (0.01) |
| | STPath as released | 0.03 (0.08) | 0.04 (0.10) | 0.04 (0.11) | 0.03 (0.07) |
| | SQUALL fine-tuned | 0.02 (0.04) | 0.02 (0.06) | 0.03 (0.07) | 0.02 (0.05) |
| | STPath fine-tuned | 0.04 (0.10) | 0.04 (0.11) | 0.05 (0.13) | 0.03 (0.09) |
| | DeepSpot-M fine-tuned | **0.05 (0.12)** | **0.05 (0.14)** | **0.05 (0.15)** | **0.04 (0.11)** |

**Supplementary Table S6. Accuracy on the most spatially autocorrelated genes.** Mean Pearson correlation of the $K$ genes with the highest mean Moran's $I$ in the training biopsies of each split, among the 15 542 genes predicted by every model, on the held-out biopsies of Supplementary Table S3 (mean over five splits), against individual-spot expression and, in parentheses, against reference expression (Supplementary Methods). Models and bold as in Supplementary Table S3. DM-R, diabetes mellitus resilient.

| Genes ($K$) | Model | AKI | CKD | DM-R | Reference |
|---|---|---|---|---|---|
| 100 | DeepSpot-M as released | 0.07 (0.13) | 0.08 (0.15) | 0.08 (0.17) | 0.05 (0.08) |
| | SQUALL as released | 0.00 (0.00) | 0.00 (0.00) | 0.01 (0.02) | 0.01 (0.02) |
| | STPath as released | 0.19 (0.34) | 0.22 (0.39) | 0.23 (0.42) | 0.19 (0.34) |
| | SQUALL fine-tuned | 0.01 (0.04) | 0.02 (0.05) | 0.02 (0.04) | 0.02 (0.04) |
| | STPath fine-tuned | 0.21 (0.38) | 0.24 (0.43) | 0.26 (0.46) | 0.21 (0.38) |
| | DeepSpot-M fine-tuned | **0.27 (0.49)** | **0.30 (0.53)** | **0.32 (0.58)** | **0.26 (0.47)** |
| 1 000 | DeepSpot-M as released | 0.04 (0.10) | 0.05 (0.12) | 0.05 (0.14) | 0.03 (0.07) |
| | SQUALL as released | 0.00 (0.00) | 0.01 (0.01) | 0.01 (0.03) | 0.01 (0.02) |
| | STPath as released | 0.12 (0.26) | 0.13 (0.29) | 0.14 (0.33) | 0.11 (0.24) |
| | SQUALL fine-tuned | 0.03 (0.07) | 0.04 (0.11) | 0.04 (0.10) | 0.04 (0.07) |
| | STPath fine-tuned | 0.14 (0.30) | 0.15 (0.33) | 0.17 (0.37) | 0.13 (0.28) |
| | DeepSpot-M fine-tuned | **0.16 (0.36)** | **0.18 (0.39)** | **0.19 (0.42)** | **0.15 (0.33)** |
| 5 000 | DeepSpot-M as released | 0.03 (0.07) | 0.04 (0.10) | 0.04 (0.10) | 0.02 (0.05) |
| | SQUALL as released | 0.00 (−0.01) | 0.01 (0.01) | 0.01 (0.02) | 0.01 (0.01) |
| | STPath as released | 0.06 (0.15) | 0.07 (0.18) | 0.08 (0.20) | 0.06 (0.14) |
| | SQUALL fine-tuned | 0.03 (0.07) | 0.04 (0.10) | 0.04 (0.11) | 0.03 (0.07) |
| | STPath fine-tuned | 0.08 (0.18) | 0.08 (0.20) | 0.09 (0.23) | 0.07 (0.16) |
| | DeepSpot-M fine-tuned | **0.09 (0.22)** | **0.10 (0.25)** | **0.10 (0.26)** | **0.08 (0.20)** |
| All | DeepSpot-M as released | 0.02 (0.04) | 0.02 (0.06) | 0.02 (0.07) | 0.02 (0.03) |
| | SQUALL as released | 0.00 (0.00) | 0.00 (0.01) | 0.01 (0.02) | 0.00 (0.01) |
| | STPath as released | 0.03 (0.08) | 0.04 (0.10) | 0.04 (0.11) | 0.03 (0.07) |
| | SQUALL fine-tuned | 0.02 (0.04) | 0.02 (0.06) | 0.03 (0.07) | 0.02 (0.05) |
| | STPath fine-tuned | 0.04 (0.10) | 0.04 (0.11) | 0.05 (0.13) | 0.03 (0.09) |
| | DeepSpot-M fine-tuned | **0.05 (0.12)** | **0.05 (0.14)** | **0.05 (0.15)** | **0.04 (0.11)** |

**Supplementary Table S7. Accuracy of the program genes.** The 103 genes of the 18 gene sets of Supplementary Table S2, each listed once, for DeepSpot-M Kidney on the held-out biopsies of the five participant-level splits. Detected, share of measured Visium spots with at least one count; within, mean Pearson correlation of virtual and reference expression across the spots of each biopsy (the metric of Figure 2b); between, Pearson correlation of virtual and measured pseudo-bulk expression across the held-out biopsies of a split, mean over splits. IFN, interferon; PT, proximal tubule; TAL, thick ascending limb; TLS, tertiary lymphoid structure.

| Gene | Detected | Pearson *r* | |
|---|---|---|---|
| | (% spots) | Within | Between |
| *Proximal tubule* | | | |
| *ALDOB* | 69.3 | 0.61 | 0.67 |
| *ACSM2A* | 48.2 | 0.59 | 0.64 |
| *GATM* | 76.6 | 0.59 | 0.67 |
| *LRP2* | 62.9 | 0.58 | 0.64 |
| *CUBN* | 47.9 | 0.53 | 0.67 |
| *SLC34A1* | 41.6 | 0.51 | 0.64 |
| *PLG* | 34.5 | 0.51 | 0.70 |
| *GSTA1* | 38.5 | 0.49 | 0.69 |
| *SLC5A2* | 29.6 | 0.44 | 0.54 |
| *Healthy PT* | | | |
| *GPX3* | 94.8 | 0.65 | 0.59 |
| *PDZK1IP1* | 75.1 | 0.56 | 0.65 |
| *APOE* | 75.6 | 0.51 | 0.61 |
| *MT1G* | 68.3 | 0.44 | 0.54 |
| *THY1* | 61.4 | 0.29 | 0.49 |
| *PT gluconeogenesis* | | | |
| *PCK1* | 55.0 | 0.53 | 0.54 |
| *FBP1* | 52.3 | 0.50 | 0.67 |
| *G6PC1* | 14.6 | 0.36 | 0.56 |
| *PC* | 19.5 | 0.24 | 0.62 |
| *Podocyte* | | | |
| *PODXL* | 60.4 | 0.52 | 0.58 |
| *NPHS2* | 26.0 | 0.51 | 0.48 |
| *WT1* | 9.2 | 0.36 | 0.28 |
| *NPHS1* | 3.0 | 0.34 | −0.24 |
| *Loop of Henle, TAL* | | | |
| *UMOD* | 89.3 | 0.48 | 0.48 |
| *EGF* | 33.9 | 0.46 | 0.68 |
| *SLC12A1* | 79.3 | 0.45 | 0.50 |
| *Distal tubule, collecting duct* | | | |
| *DEFB1* | 91.5 | 0.58 | 0.61 |
| *AQP2* | 73.9 | 0.39 | 0.48 |
| *CALB1* | 53.3 | 0.34 | 0.64 |
| *Vessel, smooth muscle* | | | |
| *VIM* | 81.5 | 0.66 | 0.77 |
| *TAGLN* | 75.1 | 0.63 | 0.56 |
| *MYH11* | 24.9 | 0.55 | 0.67 |
| *ACTA2* | 65.9 | 0.54 | 0.49 |
| *A2M* | 93.5 | 0.53 | 0.66 |
| *AQP1* | 78.0 | 0.49 | 0.55 |
| *PDGFRB* | 49.7 | 0.48 | 0.55 |
| *Fibroblast* | | | |
| *DCN* | 84.5 | 0.56 | 0.62 |
| *FBLN1* | 60.4 | 0.44 | 0.58 |
| *POSTN* | 27.7 | 0.44 | 0.44 |
| *COL1A2* | 68.8 | 0.43 | 0.59 |
| *C7* | 92.5 | 0.41 | 0.50 |
| *LUM* | 71.5 | 0.38 | 0.70 |
| *COL3A1* | 53.3 | 0.37 | 0.65 |
| *PDGFRA* | 38.1 | 0.33 | 0.63 |
| *COL1A1* | 42.7 | 0.33 | 0.61 |
| *FAP* | 4.0 | 0.07 | 0.27 |
| *Myofibroblast* | | | |
| *SPARC* | 81.9 | 0.55 | 0.58 |
| *TNC* | 41.5 | 0.37 | 0.70 |
| *INHBA* | 6.9 | 0.15 | 0.57 |
| *Inflammatory fibroblast* | | | |
| *CHI3L1* | 11.9 | 0.36 | 0.17 |
| *PXDN* | 17.1 | 0.21 | 0.64 |
| *OSMR* | 20.7 | 0.20 | 0.59 |
| *SOX4* | 37.4 | 0.17 | 0.62 |
| *IL1R1* | 27.6 | 0.15 | 0.54 |
| *MID1* | 11.1 | 0.10 | 0.39 |
| *Injury, repair PT* | | | |
| *VCAM1* | 46.0 | 0.28 | 0.66 |
| *CDH6* | 35.1 | 0.26 | 0.70 |
| *MMP7* | 49.9 | 0.26 | 0.74 |
| *CDKN1A* | 25.9 | 0.13 | 0.26 |
| *HAVCR1* | 0.4 | 0.01 | 0.29 |
| *Injury, repair (other)* | | | |
| *CD24* | 91.2 | 0.60 | 0.54 |
| *IGFBP7* | 98.3 | 0.50 | 0.60 |
| *TIMP1* | 65.8 | 0.49 | 0.69 |
| *SPP1* | 92.3 | 0.46 | 0.58 |
| *FN1* | 50.8 | 0.42 | 0.52 |
| *CST3* | 89.6 | 0.40 | 0.68 |
| *TPM1* | 72.8 | 0.37 | 0.59 |
| *DCDC2* | 37.1 | 0.32 | 0.59 |
| *PROM1* | 38.1 | 0.30 | 0.74 |
| *CLU* | 79.6 | 0.29 | 0.59 |
| *ITGB8* | 41.3 | 0.29 | 0.61 |
| *ITGB6* | 30.1 | 0.16 | 0.64 |
| *LCN2* | 10.9 | 0.12 | 0.66 |
| *SOX9* | 14.4 | 0.09 | 0.60 |
| *Adaptive PT* | | | |
| *ROBO2* | 3.0 | 0.13 | 0.08 |
| *Adaptive TAL* | | | |
| *ITGA3* | 50.8 | 0.39 | 0.57 |
| *CREB5* | 9.6 | 0.09 | 0.60 |
| *TMPRSS4* | 11.7 | 0.06 | 0.41 |
| *Degenerative* | | | |
| *GDF15* | 33.3 | 0.27 | 0.35 |
| *NAMPT* | 30.2 | 0.13 | 0.43 |
| *CDKN2A* | 0.9 | 0.06 | 0.26 |
| *Immune* | | | |
| *B2M* | 97.5 | 0.52 | 0.60 |
| *C3* | 33.6 | 0.27 | 0.68 |
| *CD68* | 31.3 | 0.24 | 0.70 |
| *MRC1* | 21.7 | 0.23 | 0.67 |
| *CCL2* | 37.5 | 0.22 | 0.60 |
| *PTPRC* | 19.4 | 0.20 | 0.73 |
| *IFN-γ, CXCL9* | | | |
| *HLA-DRA* | 85.3 | 0.47 | 0.71 |
| *STAT1* | 43.8 | 0.24 | 0.57 |
| *CD3E* | 7.9 | 0.18 | 0.70 |
| *CD8A* | 4.7 | 0.13 | 0.58 |
| *CXCL9* | 5.6 | 0.11 | 0.32 |
| *TNF* | 2.6 | 0.07 | 0.38 |
| *CXCL10* | 5.9 | 0.06 | 0.30 |
| *IFNG* | 0.2 | 0.03 | 0.05 |
| *B cell, TLS* | | | |
| *CCL21* | 38.1 | 0.31 | 0.54 |
| *CCL19* | 18.4 | 0.28 | 0.59 |
| *LTB* | 18.0 | 0.26 | 0.70 |
| *MZB1* | 12.0 | 0.17 | 0.60 |
| *JCHAIN* | 36.4 | 0.16 | 0.38 |
| *CD79A* | 5.7 | 0.16 | 0.66 |
| *MS4A1* | 3.4 | 0.14 | 0.49 |
| *CD19* | 1.2 | 0.10 | 0.47 |
| *CXCL13* | 0.5 | 0.09 | 0.26 |

**Supplementary Table S8. Virtual programs and eGFR beyond histopathology.** Rank partial correlation (FDR) of participant-level atlas program scores of FFPE slides with baseline eGFR in the 231 participants without AKI with pathologist scores of interstitial fibrosis (IF) and tubular atrophy (TA) (the partial correlation of 0.32 in the Results covers all 439 FFPE participants without AKI), adjusted for site, sex, age, and scanner (covariates), additionally for IF and TA, and additionally for the adjudicated diagnosis. FDR was controlled over the ten programs per column. In a linear model of eGFR with all these variables, eGFR was 5.7 ml/min per 1.73 $m^2$ higher per SD of the proximal tubule program score (95% CI, 2.2 to 9.3). In repeated cross-validation without diagnosis (Supplementary Methods), adding the 18 virtual program scores to the covariates, IF, and TA increased the explained variance ($R^2$) of eGFR from 0.35 to 0.41. PT, proximal tubule; TAL, thick ascending limb; TLS, tertiary lymphoid structure.

| Tissue (*n*) | Program | Covariates | + IF, TA | + IF, TA, diagnosis |
|---|---|---|---|---|
| FFPE (231) | Proximal tubule | 0.38 (<0.001) | 0.19 (0.02) | 0.18 (0.04) |
| | Healthy PT | 0.33 (<0.001) | 0.17 (0.03) | 0.16 (0.05) |
| | PT gluconeogenesis | 0.39 (<0.001) | 0.20 (0.02) | 0.19 (0.04) |
| | Loop of Henle, TAL | 0.40 (<0.001) | 0.17 (0.03) | 0.16 (0.05) |
| | Injury, repair PT | −0.17 (0.010) | 0.00 (0.95) | 0.00 (1.00) |
| | Adaptive PT | −0.28 (<0.001) | −0.13 (0.10) | −0.12 (0.17) |
| | Fibroblast | −0.26 (<0.001) | −0.08 (0.34) | −0.08 (0.38) |
| | Immune | −0.25 (<0.001) | −0.06 (0.46) | −0.06 (0.40) |
| | IFN-$\gamma$, CXCL9 | −0.26 (<0.001) | −0.06 (0.43) | −0.07 (0.40) |
| | B cell, TLS | −0.30 (<0.001) | −0.11 (0.19) | −0.11 (0.17) |

**Supplementary Table S9. Withheld Xenium panel genes by resolution, enrollment group, and Xenium panel.** Pearson correlation of virtual with measured expression of the 233 panel genes on chromosomes 1, 3, 5, 7, and 9, which were withheld from the adaptation of DeepSpot-M Kidney to the Xenium sections, before and after adaptation, given as the median over sections of the median over genes. Cell, adaptation and scoring on single cells (Figure 4g); 56 µm patch, adaptation and scoring on the cells pooled in the 56 µm lattice of the atlas. Values are against the expression of individual cells or patches and, in parentheses, against reference expression (the metric of Figure 4g). Improved, sections with a higher correlation after adaptation. The sections of participants outside the Visium training are those of Figure 4g. AKI, acute kidney injury; CKD, chronic kidney disease; DM-R, diabetes mellitus resilient.

| Resolution | Participants | Subgroup | Sections | Before | After | Improved |
|---|---|---|---|---|---|---|
| Cell | Outside Visium training | All | 34 | 0.04 (0.11) | 0.09 (0.21) | 34 (34) |
| | | Xenium hKidney panel | 17 | 0.02 (0.08) | 0.09 (0.22) | 17 (17) |
| | | Xenium hMulti panel | 17 | 0.05 (0.13) | 0.09 (0.20) | 17 (17) |
| | | AKI | 1 | 0.02 (0.06) | 0.09 (0.21) | 1 (1) |
| | | CKD | 25 | 0.04 (0.13) | 0.09 (0.21) | 25 (25) |
| | | DM-R | 7 | 0.02 (0.07) | 0.09 (0.18) | 7 (7) |
| | | Reference | 1 | 0.00 (0.03) | 0.07 (0.17) | 1 (1) |
| | All | All | 49 | 0.04 (0.10) | 0.09 (0.21) | 49 (48) |
| | | AKI | 6 | 0.04 (0.12) | 0.10 (0.23) | 6 (6) |
| | | CKD | 31 | 0.04 (0.12) | 0.09 (0.21) | 31 (31) |
| | | DM-R | 8 | 0.02 (0.07) | 0.08 (0.18) | 8 (8) |
| | | Reference | 4 | 0.01 (0.04) | 0.06 (0.15) | 4 (3) |
| 56 µm patch | Outside Visium training | All | 34 | 0.12 (0.23) | 0.24 (0.41) | 34 (34) |
| | | Xenium hKidney panel | 17 | 0.10 (0.21) | 0.18 (0.35) | 17 (17) |
| | | Xenium hMulti panel | 17 | 0.14 (0.30) | 0.28 (0.46) | 17 (17) |
| | | AKI | 1 | 0.14 (0.26) | 0.18 (0.35) | 1 (1) |
| | | CKD | 25 | 0.13 (0.23) | 0.26 (0.43) | 25 (25) |
| | | DM-R | 7 | 0.09 (0.19) | 0.17 (0.28) | 7 (7) |
| | | Reference | 1 | 0.07 (0.18) | 0.18 (0.44) | 1 (1) |
| | All | All | 49 | 0.12 (0.24) | 0.23 (0.39) | 47 (46) |
| | | AKI | 6 | 0.15 (0.29) | 0.17 (0.29) | 4 (4) |
| | | CKD | 31 | 0.13 (0.29) | 0.26 (0.42) | 31 (31) |
| | | DM-R | 8 | 0.09 (0.20) | 0.16 (0.28) | 8 (7) |
| | | Reference | 4 | 0.09 (0.21) | 0.19 (0.37) | 4 (4) |

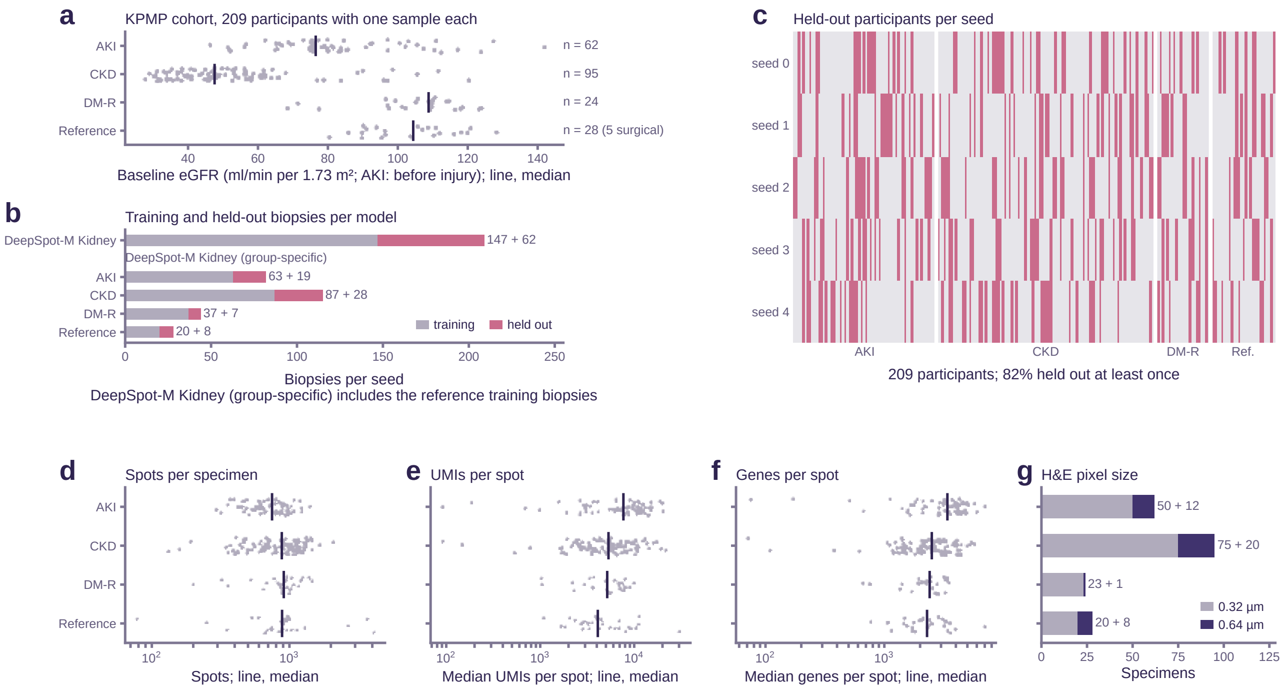


**Supplementary Figure S1. Cohort, participant-level splits, and Visium data.** (a) Baseline eGFR by enrollment group (AKI, before injury); line, median. (b) Training and held-out biopsies per split for DeepSpot-M Kidney and DeepSpot-M Kidney (group-specific). (c) Participants held out in each of the five splits (pink); 171 of 209 (82%) were held out at least once. (d–g) Spots, median UMIs and median genes per spot (logarithmic scale; line, median), and H&E pixel size per specimen by group (Supplementary Table S1). AKI, acute kidney injury; CKD, chronic kidney disease; DM-R, diabetes mellitus resilient; KPMP, Kidney Precision Medicine Project; Ref., reference; UMI, unique molecular identifier.

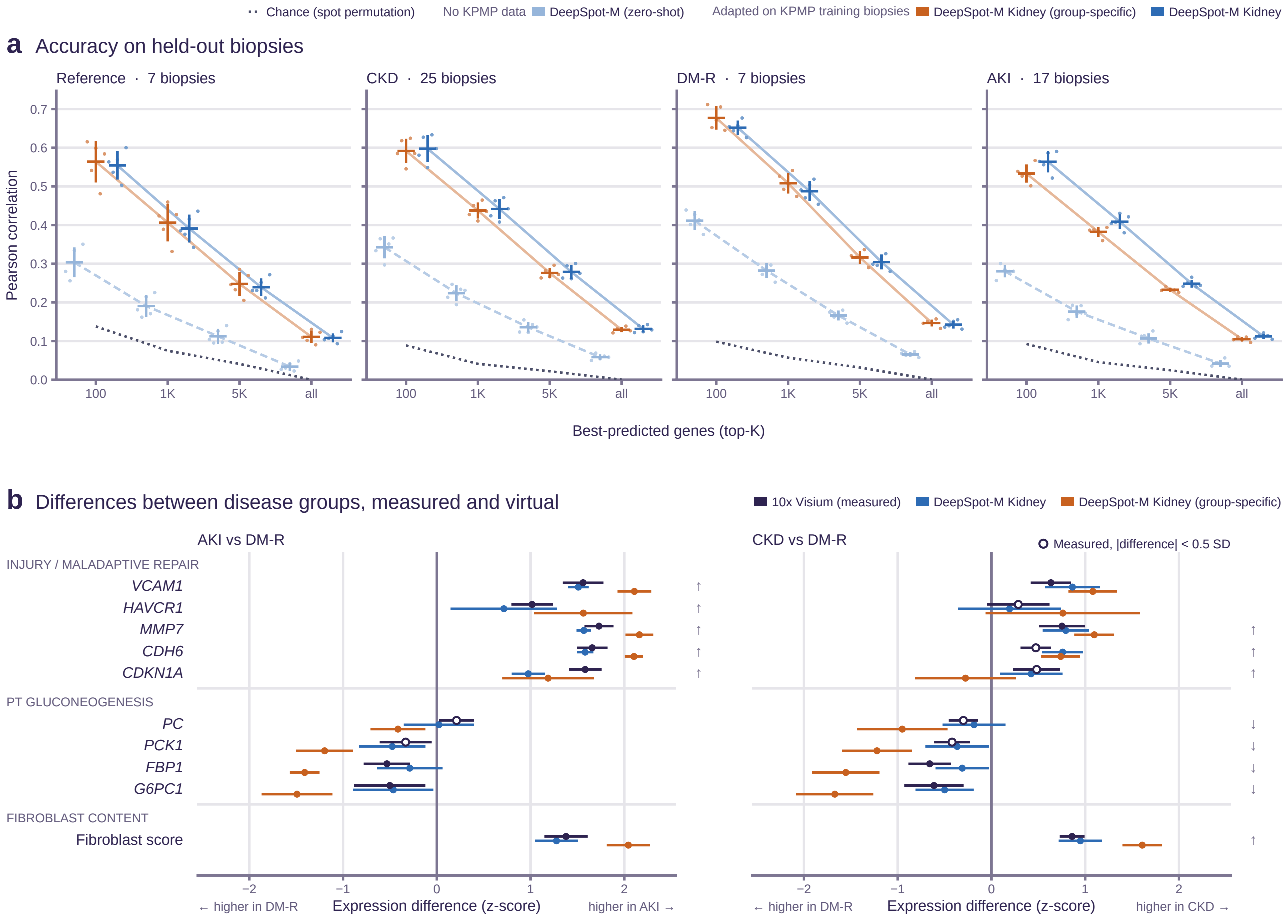


**Supplementary Figure S2. DeepSpot-M Kidney and the group-specific models.** DeepSpot-M (zero-shot) had no KPMP data, and DeepSpot-M Kidney (all enrollment groups) and DeepSpot-M Kidney (group-specific) (one model per enrollment group) were adapted on the training biopsies of each split. (a) Accuracy of the $K$ best-predicted genes on the held-out biopsies, as in Figure 2a. (b) Differences between disease groups as in Figure 2d, including DeepSpot-M Kidney (group-specific), which enlarged 12 of the 13 measured differences of at least 0.5 SD (filled measured points). AKI, acute kidney injury; CKD, chronic kidney disease; DM-R, diabetes mellitus resilient; KPMP, Kidney Precision Medicine Project; PT, proximal tubule.

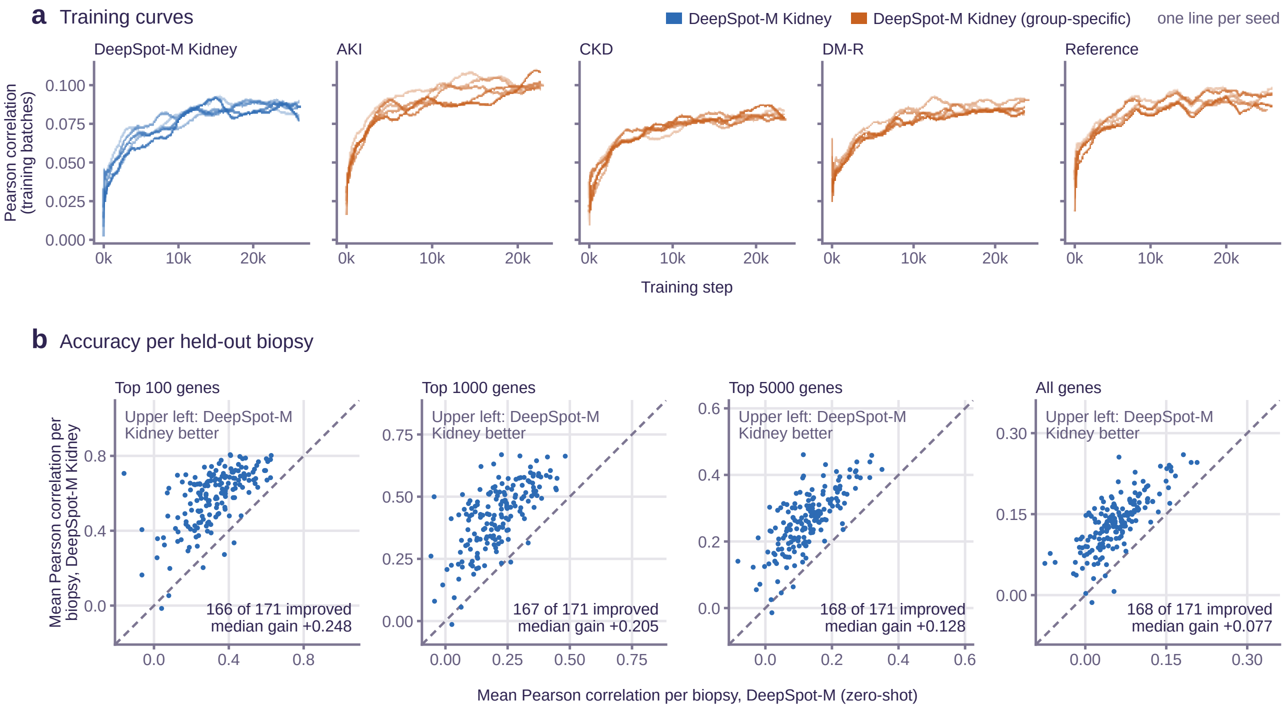


**Supplementary Figure S3. Training curves and per-biopsy accuracy.** (a) Pearson correlation on training batches during adaptation of DeepSpot-M Kidney and DeepSpot-M Kidney (group-specific), one line per split. (b) Mean Pearson correlation with reference expression per held-out biopsy of DeepSpot-M (zero-shot) and DeepSpot-M Kidney for each model's 100, 1 000, and 5 000 best-predicted genes and all genes, averaged over the splits that held the biopsy out; dashed line, equal accuracy. AKI, acute kidney injury; CKD, chronic kidney disease; DM-R, diabetes mellitus resilient.

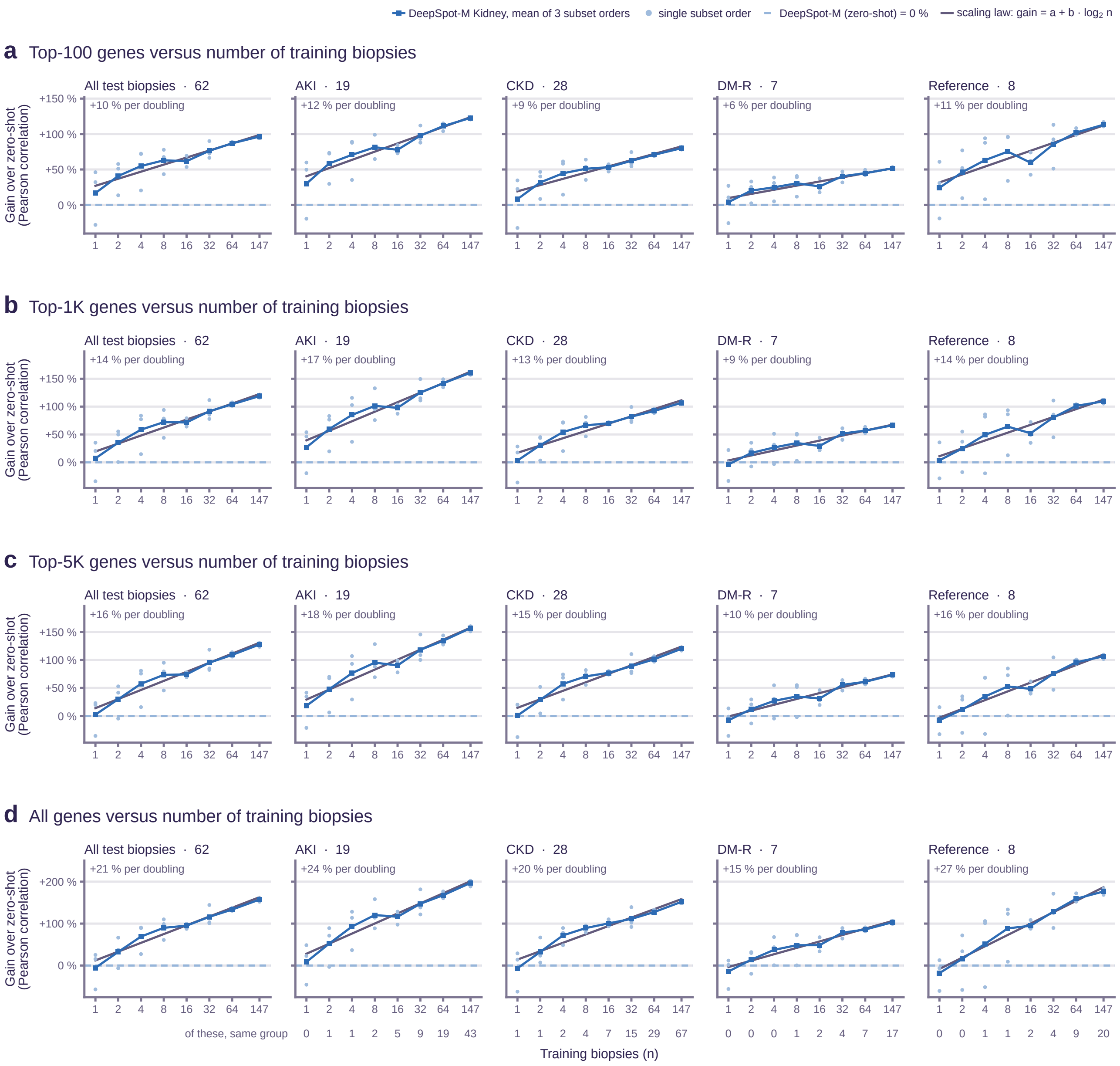


**Supplementary Figure S4. Accuracy as a function of the number of training biopsies.** DeepSpot-M Kidney adapted on nested, group-stratified subsets of *n* of the 147 training biopsies of split 0 and evaluated on its 62 held-out biopsies, overall and per group. Gain in mean Pearson correlation over DeepSpot-M (zero-shot) (dashed line, 0%) for the (a) 100, (b) 1 000, and (c) 5 000 best-predicted genes and (d) all genes. Squares, mean of three subset orders; dots, single orders; gray line, fitted scaling law (gain per doubling above each panel); numbers below the axis, training biopsies of the same group. AKI, acute kidney injury; CKD, chronic kidney disease; DM-R, diabetes mellitus resilient.

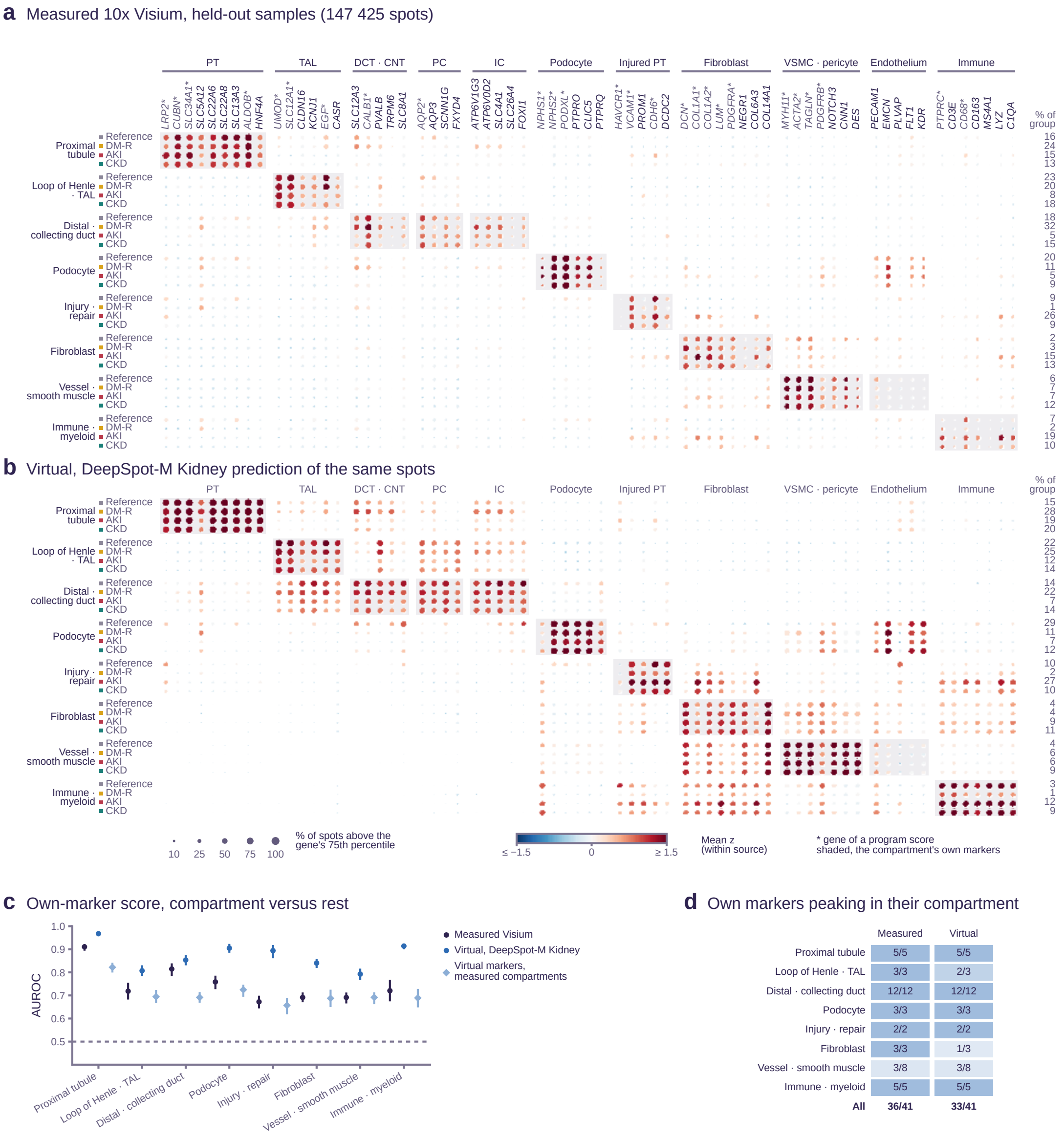


**Supplementary Figure S5. Marker genes of program compartments in measured and virtual profiles.** Each spot of the 171 held-out biopsies was assigned to its highest-scoring program (Supplementary Methods); rows, program compartments by disease group. (a) Measured 10x Genomics Visium expression. (b) Virtual expression of the same spots (DeepSpot-M Kidney model that did not train on the biopsy). Columns, marker genes by cell type; dot size, percentage of spots above the gene's 75th percentile; color, mean $z$-score; asterisk, program-score gene; shading, the row compartment's markers; right, percentage of the group's spots in the compartment. (c) AUROC of each compartment's mean marker $z$-score (markers outside the program gene sets), its spots versus all others; bars, 95% bootstrap CI over biopsies; light diamonds, virtual scores on compartments assigned from measured expression. (d) Markers outside the program gene sets whose highest mean $z$-score fell in their own compartment. AKI, acute kidney injury; AUROC, area under the receiver operating characteristic curve; CI, confidence interval; CKD, chronic kidney disease; CNT, connecting tubule; DCT, distal convoluted tubule; DM-R, diabetes mellitus resilient; IC, intercalated cell; PC, principal cell; PT, proximal tubule; TAL, thick ascending limb; VSMC, vascular smooth muscle cell.

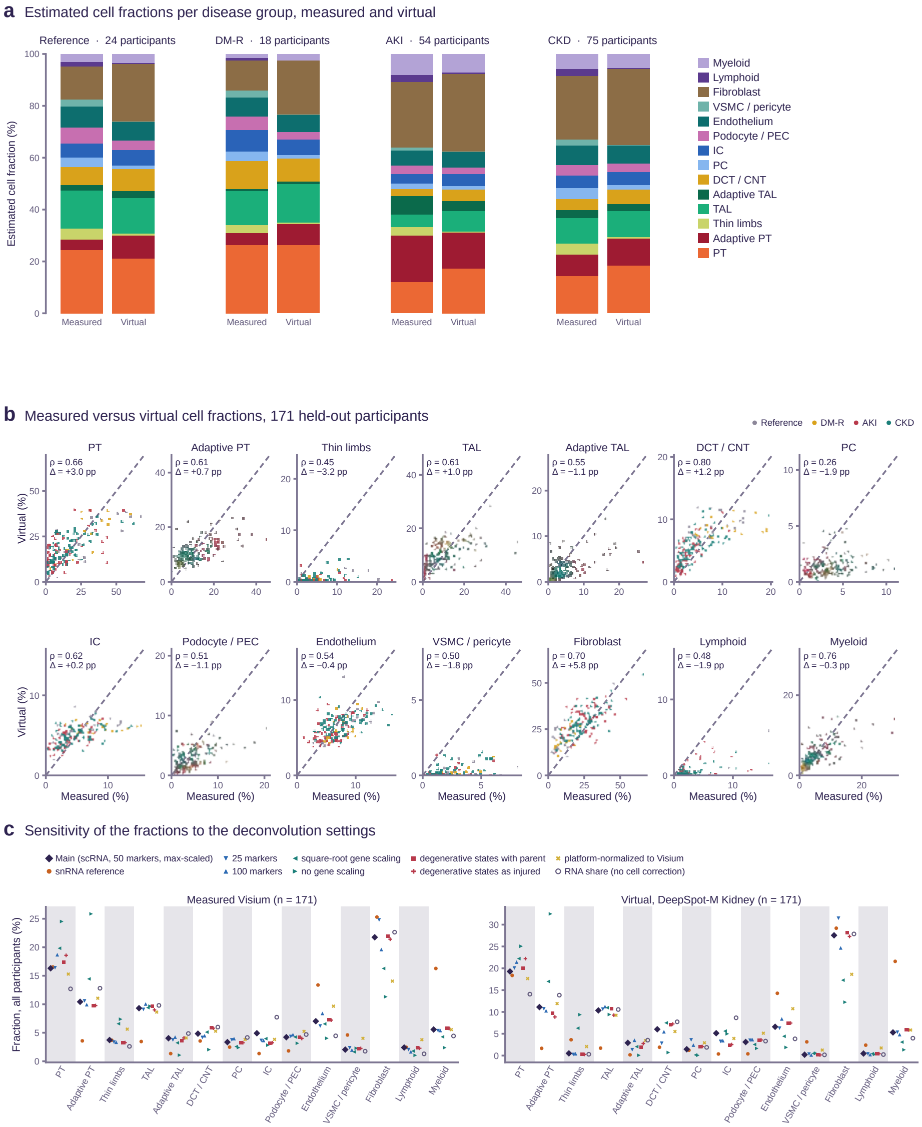


**Supplementary Figure S6. Cell type fractions estimated by deconvolution.** Measured 10x Genomics Visium spots of the 171 held-out participants and their virtual expression from the DeepSpot-M Kidney model that did not train on the participant, deconvolved with the KPMP single-cell reference (Supplementary Methods). (a) Mean cell type fractions per disease group. (b) Measured versus virtual fraction per cell type; dots, participants; $\rho$, Spearman correlation; $\Delta$, mean difference (virtual minus measured) in percentage points (pp); dashed line, identity. Virtual fractions underestimated adaptive PT and TAL cells in AKI and overestimated fibroblasts in biopsies with little fibrosis. (c) Mean fraction over all participants under alternative deconvolution settings. AKI, acute kidney injury; CKD, chronic kidney disease; CNT, connecting tubule; DCT, distal convoluted tubule; DM-R, diabetes mellitus resilient; IC, intercalated cell; KPMP, Kidney Precision Medicine Project; PC, principal cell; PEC, parietal epithelial cell; PT, proximal tubule; TAL, thick ascending limb; VSMC, vascular smooth muscle cell.

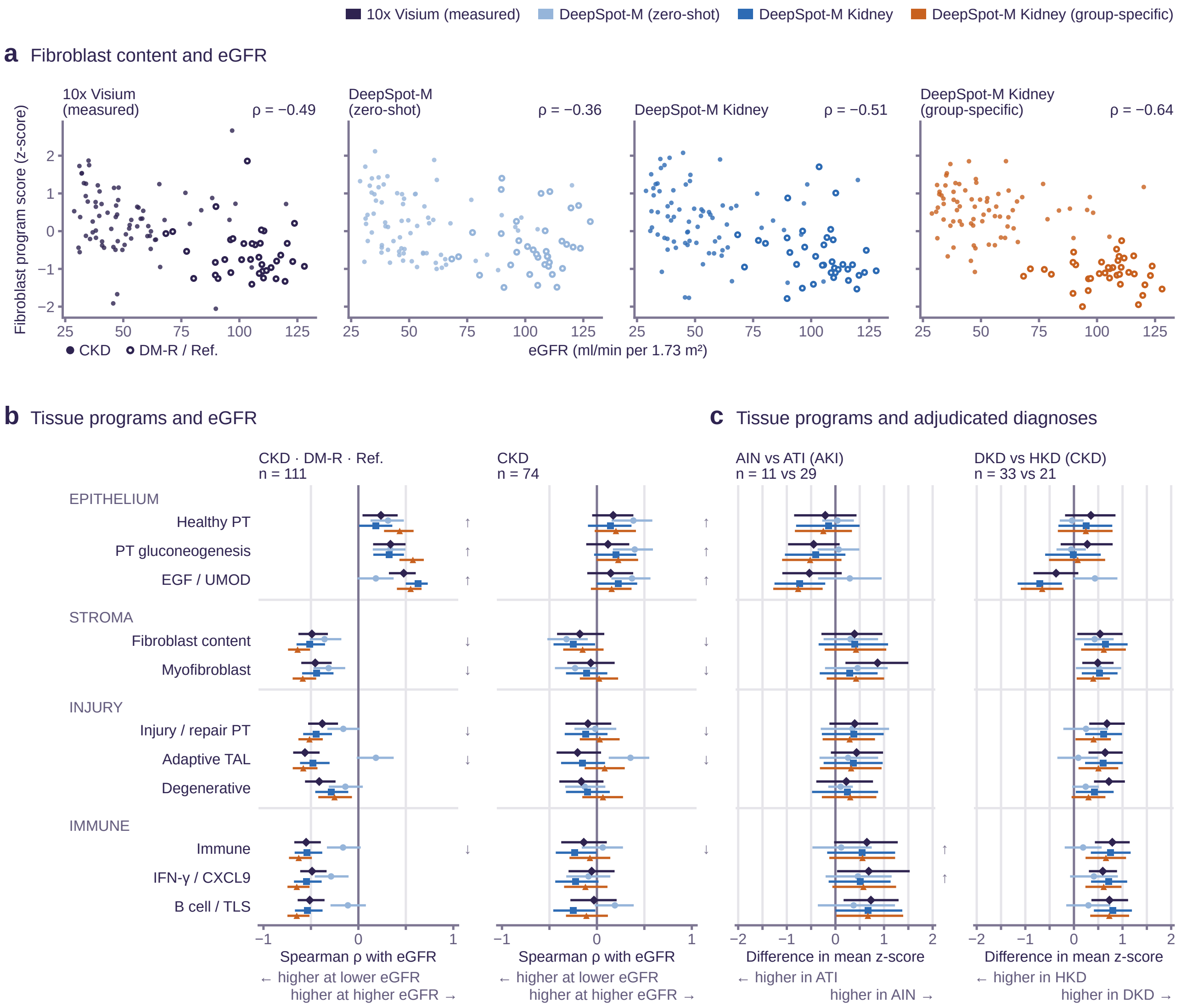


**Supplementary Figure S7. Virtual tissue programs, kidney function, and adjudicated diagnoses.** Each of the 171 held-out biopsies was scored only by models that had not seen it. (a) Fibroblast program score versus baseline eGFR in participants with CKD, DM-R, or reference tissue ($n$ = 111); $\rho$, Spearman correlation. (b) Spearman correlation of program scores with eGFR in the same participants and in CKD alone ($n$ = 74). (c) Difference in mean program score between AIN and ATI ($n$ = 11 versus 29) and between DKD and HKD ($n$ = 33 versus 21). Bars, 95% bootstrap CI; arrows, direction of measured associations whose CI excluded zero. AIN, acute interstitial nephritis; AKI, acute kidney injury; ATI, acute tubular injury; CI, confidence interval; CKD, chronic kidney disease; DKD, diabetic kidney disease; DM-R, diabetes mellitus resilient; EGF, epidermal growth factor; HKD, hypertensive kidney disease; IFN, interferon; PT, proximal tubule; Ref., reference; TAL, thick ascending limb; TLS, tertiary lymphoid structure.

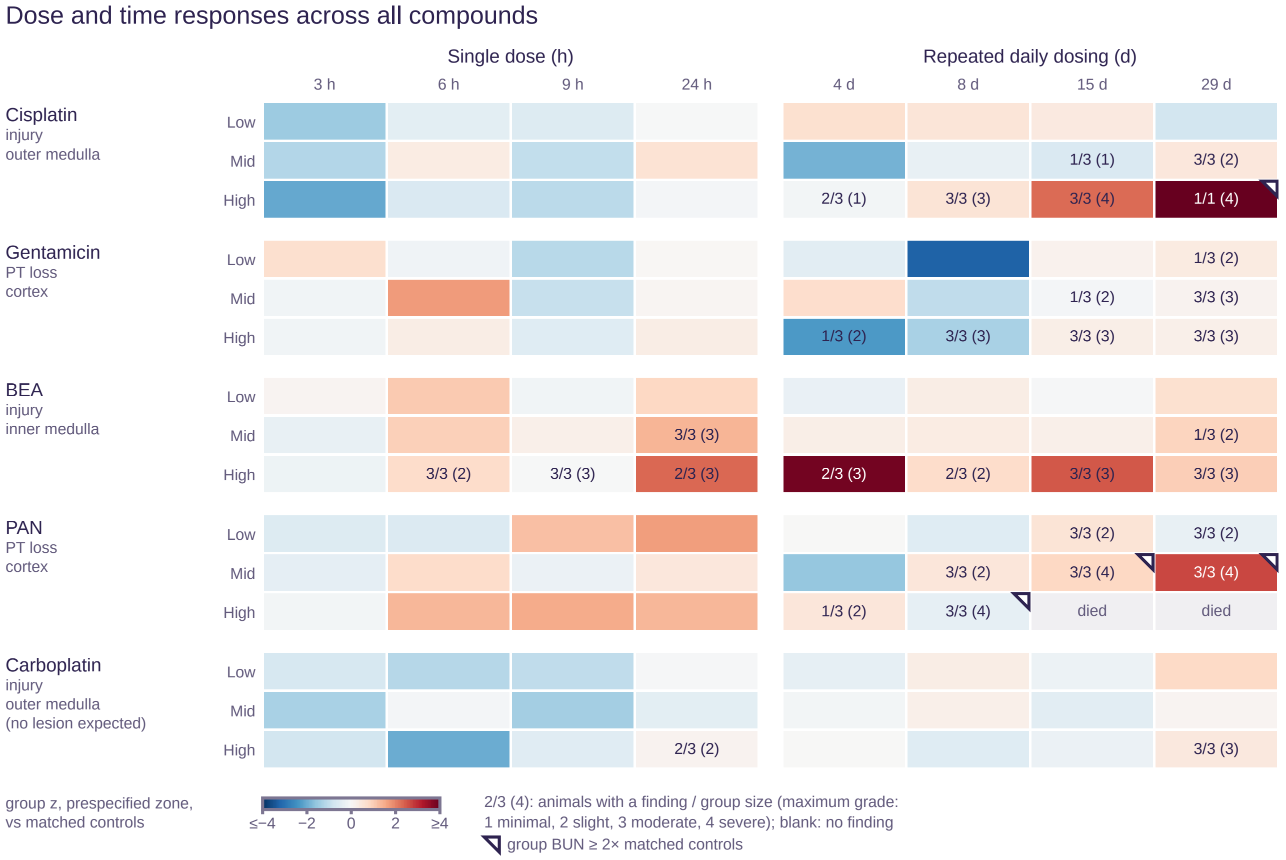


**Supplementary Figure S8. Dose and time responses for five nephrotoxic compounds in rats.** Mean standardized score of each dose group in the region of expected injury, relative to time-matched controls, after a single dose (hours) or daily dosing (days). Numbers, rats with a histopathologic finding/group size (maximum grade 1, minimal, to 4, severe); corner mark, group BUN at least twice that of matched controls. Doxorubicin is not shown because of a control-batch offset at 29 days. BEA, 2-bromoethylamine; BUN, blood urea nitrogen; PAN, puromycin aminonucleoside; PT, proximal tubule.

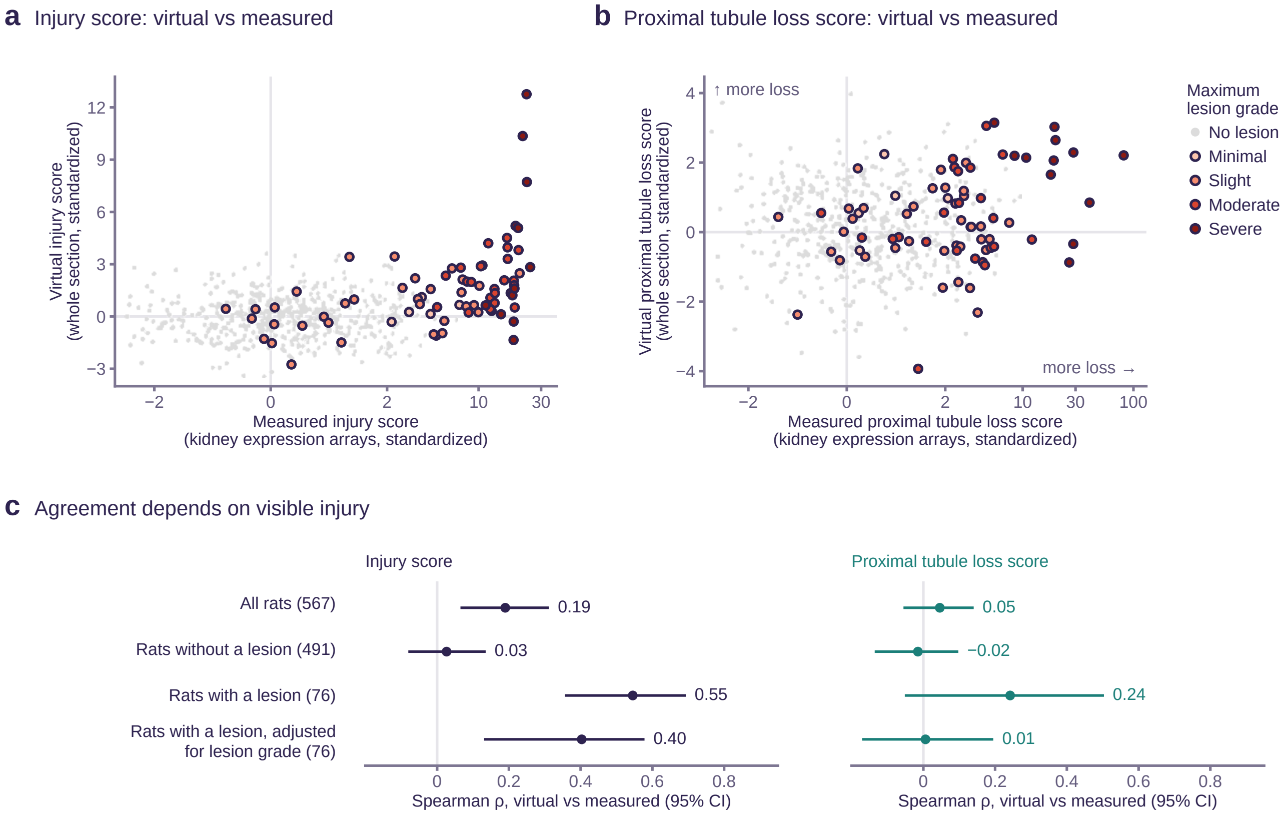


**Supplementary Figure S9. Virtual versus measured kidney transcription in rats.** Array-measured scores of separate kidney fragments of the 567 rats of Figure 3 versus virtual whole-section scores on the same genes, both standardized against time-matched controls (Supplementary Methods). (a) Injury and (b) proximal tubule loss score per rat; gray, rats without a lesion; colors, maximum lesion grade; horizontal axis linear between −2 and 2 and logarithmic beyond. (c) Spearman correlation of virtual and measured scores by group of rats; bars, 95% bootstrap CI over control blocks. CI, confidence interval.

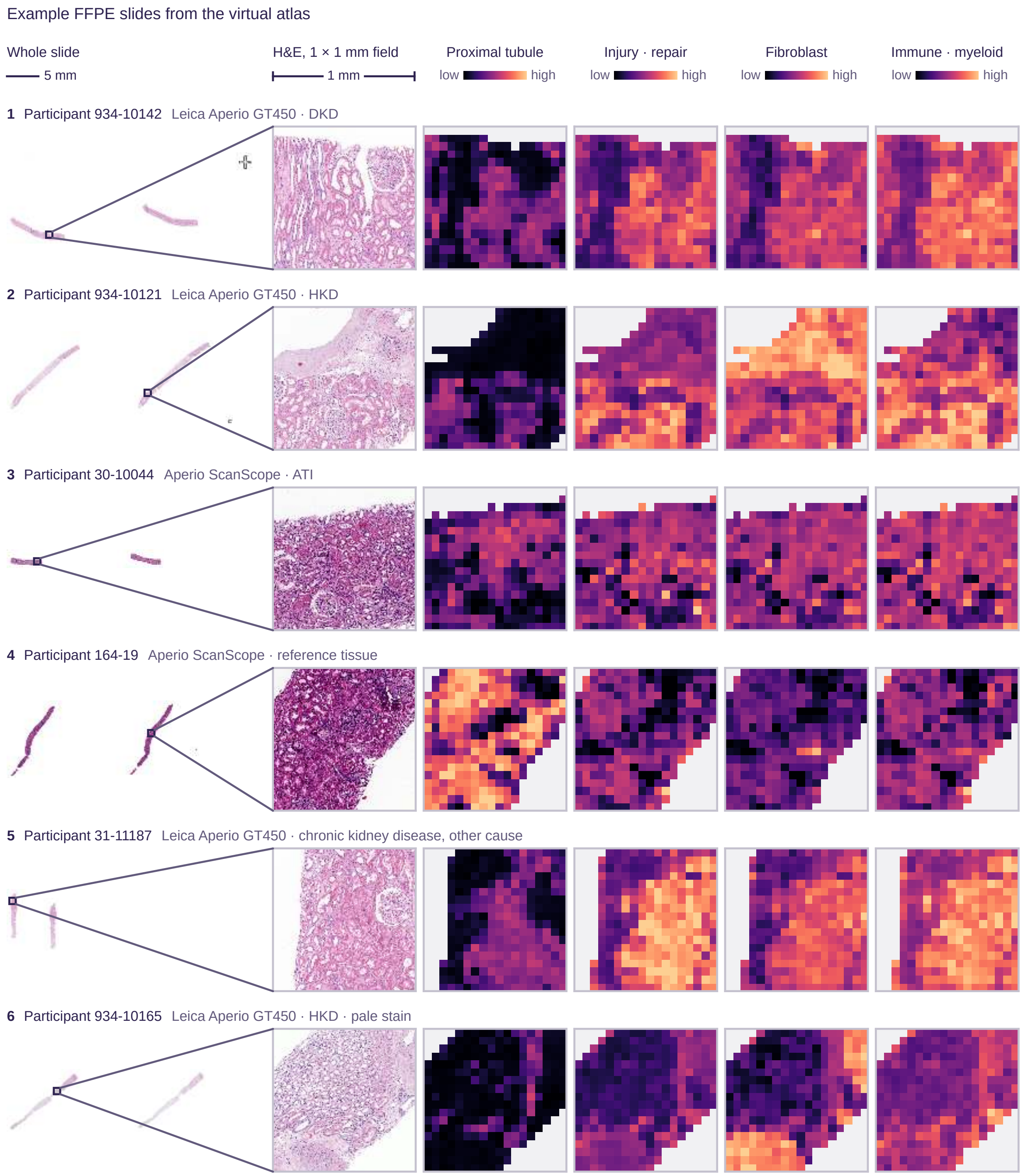


**Supplementary Figure S10. Example slides from the virtual atlas.** Six FFPE slides, one per group of scanner, diagnosis, and staining intensity. Per slide, whole-slide H&E image with the zoomed field, H&E image of that field, and virtual program scores per 56 µm virtual spot (one color scale per program). H&E images are adjusted uniformly for display. Scale bars, 5 mm (whole slides) and 1 mm (fields). ATI, acute tubular injury; DKD, diabetic kidney disease; FFPE, formalin-fixed paraffin-embedded; HKD, hypertensive kidney disease.

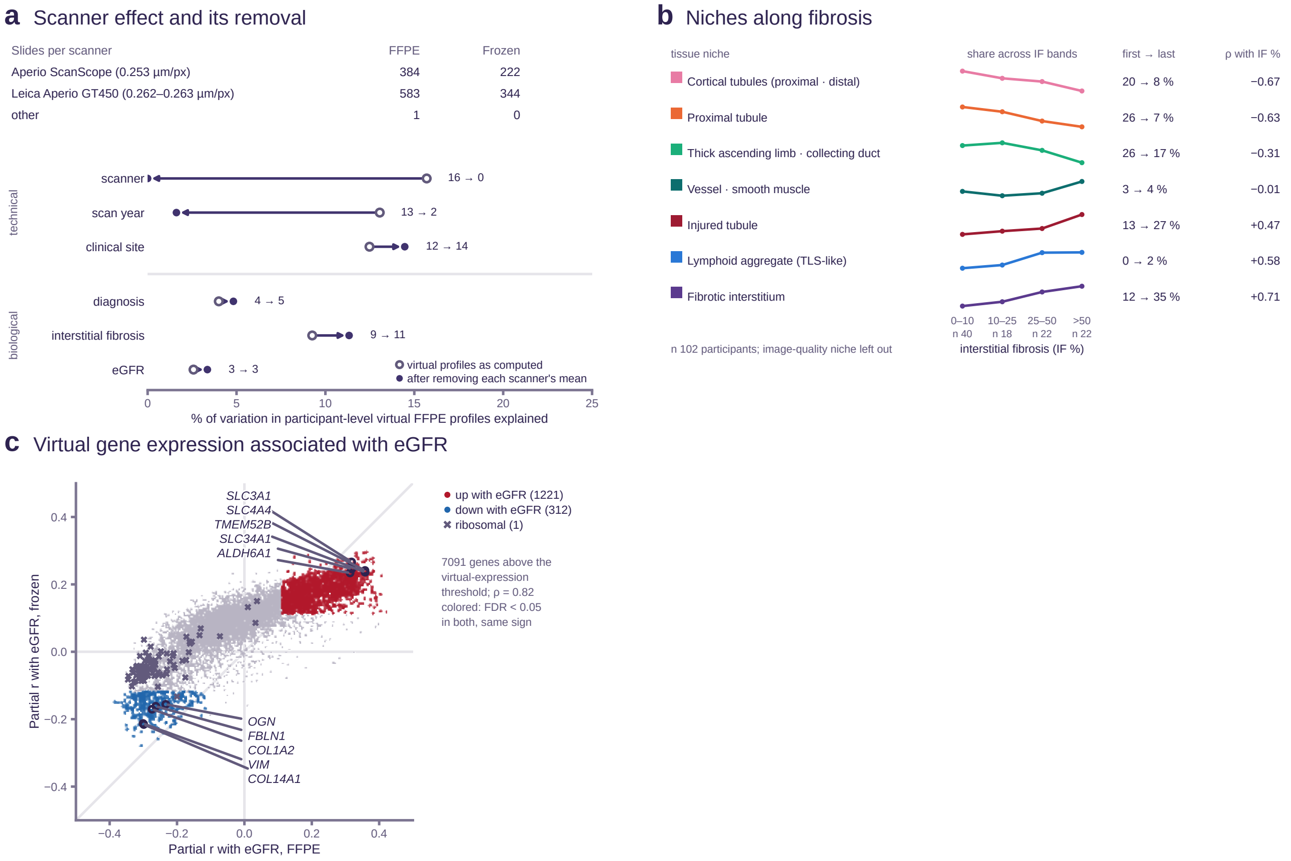


**Supplementary Figure S11. Scanner variation, niche composition, and gene-level eGFR associations in the virtual atlas.** (a) Slides per scanner and tissue preparation, and share of variation in participant-level virtual FFPE profiles explained by technical and biological factors (Supplementary Methods), before (open circles) and after (filled) removing each scanner's mean. (b) Share of each tissue niche across IF bands (image-quality niche excluded; 102 participants); numbers, shares in the first and last band and Spearman correlation with IF. (c) Partial correlation with eGFR of 7 091 genes in FFPE versus frozen slides of the same participants (Spearman $\rho$, 0.82); colors, genes at FDR $<$ 0.05 in both with the same sign (1 221 higher and 312 lower at higher eGFR, and one ribosomal gene). FDR, false discovery rate; FFPE, formalin-fixed paraffin-embedded; IF, interstitial fibrosis; TLS, tertiary lymphoid structure.

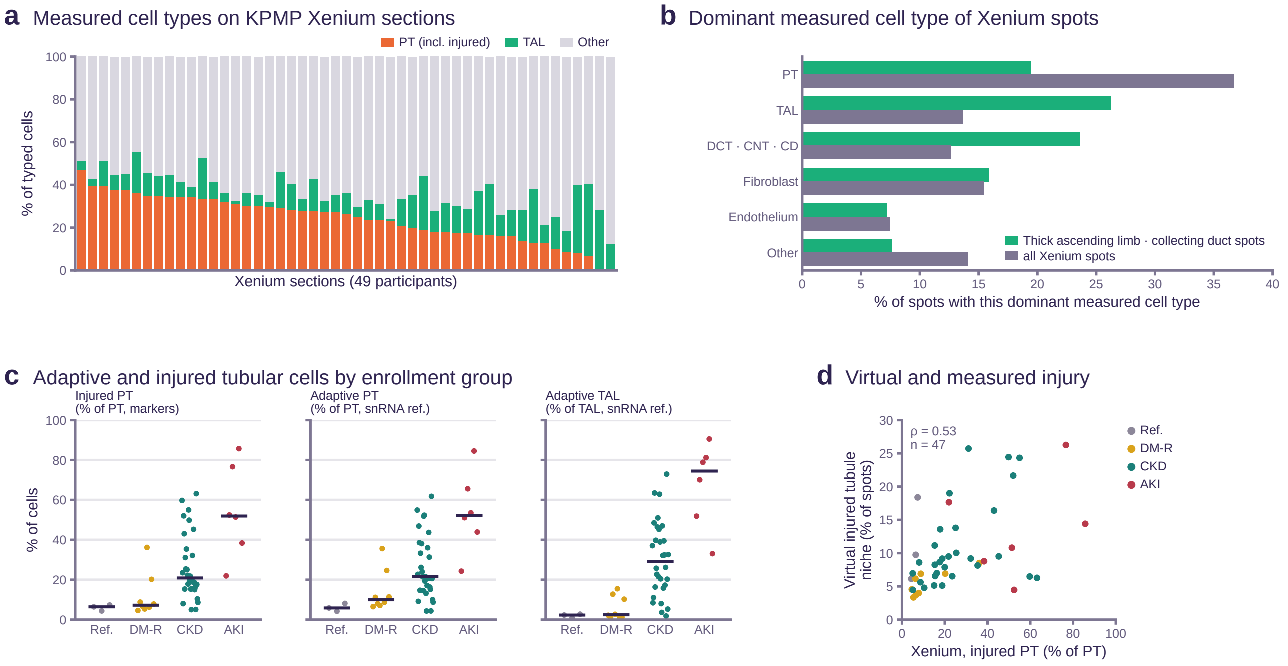


**Supplementary Figure S12. Cell types and adaptive cell states measured by Xenium.** (a) Measured cell types per section on 49 KPMP Xenium sections, sorted by PT share (including injured PT); median per section, 27% PT and 10% TAL cells. (b) Dominant measured cell type of the Xenium spots of the thick ascending limb and collecting duct niche and of all spots (Supplementary Methods). PT was 24% of typed cells but, as PT cells are large, 38% of cell area and 37% of spots by dominant type. (c) Injured PT cells (marker typing) and adaptive PT and TAL cells (transfer from the KPMP single-nucleus reference) as shares of PT or TAL cells per enrollment group, in the 47 sections with at least 200 PT cells; dots, sections; lines, median; Kruskal–Wallis $P < 0.001$ for adaptive PT and $< 0.001$ for adaptive TAL cells, with medians of 6% and 2% in reference and 52% and 74% in AKI sections. (d) Share of all spots in the virtual injured tubule niche versus share of injured PT cells among measured PT cells per section (different denominators, compared by rank); colors, enrollment group; $\rho$, Spearman correlation. AKI, acute kidney injury; CD, collecting duct; CKD, chronic kidney disease; CNT, connecting tubule; DCT, distal convoluted tubule; DM-R, diabetes mellitus resilient; KPMP, Kidney Precision Medicine Project; PT, proximal tubule; Ref., reference; TAL, thick ascending limb.

#### Supplementary References


S1. Elnaggar A, Heinzinger M, Dallago C, et al. ProtTrans: toward understanding the language of life through self-supervised learning. IEEE Transactions on Pattern Analysis and Machine Intelligence. 2022;44(10):7112-27. doi:10.1109/TPAMI.2021.3095381.

S2. Hu D, Szklarczyk D, von Mering C, Jensen LJ. SPACE: STRING proteins as complementary embeddings. Bioinformatics. 2025;41(9):btaf496. doi:10.1093/bioinformatics/btaf496.

S3. Karasikov M, van Doorn J, Känzig N, et al. Training state-of-the-art pathology foundation models with orders of magnitude less data. Medical Image Computing and Computer Assisted Intervention – MICCAI 2025, Lecture Notes in Computer Science. 2025:573-83. doi:10.1007/978-3-032-04984-1_55.

S4. Nonchev K, Dawo S, Silina K, et al. DeepSpot-M: a multimodal foundation model for transcriptome-wide virtual spatial transcriptomics from histology. medRxiv. 2026. doi:10.64898/2026.06.19.26356060.

S5. Huang T, Liu T, Babadi M, et al. STPath: a generative foundation model for integrating spatial transcriptomics and whole-slide images. NPJ Digit Med. 2025;8(1):659. doi:10.1038/s41746-025-02020-3.

S6. Zhang Z, Qin B, Zhao Y, et al. A multimodal foundation model to integrate histology with spatial molecular programs. bioRxiv. 2026. doi:10.64898/2026.06.01.729028.

S7. Xu H, Usuyama N, Bagga J, et al. A whole-slide foundation model for digital pathology from real-world data. Nature. 2024;630(8015):181–188. doi:10.1038/s41586-024-07441-w.

S8. Lake BB, Menon R, Winfree S, et al. An atlas of healthy and injured cell states and niches in the human kidney. Nature. 2023;619(7970):585–594. doi:10.1038/s41586-023-05769-3.

S9. Elosua-Bayes M, Nieto P, Mereu E, et al. SPOTlight: seeded NMF regression to deconvolute spatial transcriptomics spots with single-cell transcriptomes. Nucleic Acids Res. 2021;49(9):e50. doi:10.1093/nar/gkab043.

S10. Racle J, de Jonge K, Baumgaertner P, et al. Simultaneous enumeration of cancer and immune cell types from bulk tumor gene expression data. eLife. 2017;6:e26476. doi:10.7554/elife.26476.

S11. Igarashi Y, Nakatsu N, Yamashita T, et al. Open TG-GATEs: a large-scale toxicogenomics database. Nucleic Acids Res. 2015;43(D1):D921-7. doi:10.1093/nar/gku955.

S12. Szklarczyk D, Kirsch R, Koutrouli M, et al. The STRING database in 2023: protein–protein association networks and functional enrichment analyses for any sequenced genome of interest. Nucleic Acids Res. 2023;51(D1):D638-46. doi:10.1093/nar/gkac1000.

S13. Uehara T, Yamate J, Torii M, Maruyama T. Comparative nephrotoxicity of cisplatin and nedaplatin: mechanisms and histopathological characteristics. J Toxicol Pathol. 2011;24(2):87-94. doi:10.1293/tox.24.87.

S14. Blanco-Gozalo V, Quiros Y, Vicente-Vicente L, et al. Urinary GM2AP coincides with renal cortical damage and grades cisplatin nephrotoxicity severity in rats. Toxicology. 2024;508:153919. doi:10.1016/j.tox.2024.153919.

S15. Murray G, Wyllie RG, Hill GS, et al. Experimental papillary necrosis of the kidney. I. Morphologic and functional data. Am J Pathol. 1972;67(2):285-302.

S16. Hedlund LW, Maronpot RR, Johnson GA, et al. Magnetic resonance microscopy of toxic renal injury induced by bromoethylamine in rats. Fundam Appl Toxicol. 1991;16(4):787-97. doi:10.1016/0272-0590(91)90164-y.

S17. Wedeen RP, Batuman V, Cheeks C, et al. Transport of gentamicin in rat proximal tubule. Lab Invest. 1983;48(2):212-23.

S18. Vernier RL, Papermaster BW, Good RA. Aminonucleoside nephrosis. I. Electron microscopic study of the renal lesion in rats. J Exp Med. 1959;109(1):115-26. doi:10.1084/jem.109.1.115.

S19. Cianciolo R, Yoon L, Krull D, et al. Gene expression analysis and urinary biomarker assays reveal activation of tubulointerstitial injury pathways in a rodent model of chronic proteinuria (doxorubicin nephropathy). Nephron Exp Nephrol. 2013;124(1-2):1-10. doi:10.1159/000355542.

S20. Yasumasu T, Ueda T, Uozumi J, et al. Comparative study of cisplatin and carboplatin on pharmacokinetics, nephrotoxicity and effect on renal nuclear DNA synthesis in rats. Pharmacol Toxicol. 1992;70(2):143-7. doi:10.1111/j.1600-0773.1992.tb00445.x.

S21. Irizarry RA, Hobbs B, Collin F, et al. Exploration, normalization, and summaries of high density oligonucleotide array probe level data. Biostatistics. 2003;4(2):249-64. doi:10.1093/biostatistics/4.2.249.

S22. Korsunsky I, Millard N, Fan J, et al. Fast, sensitive and accurate integration of single-cell data with Harmony. Nat Methods. 2019;16(12):1289-96. doi:10.1038/s41592-019-0619-0.

S23. Wolf FA, Angerer P, Theis FJ. SCANPY: large-scale single-cell gene expression data analysis. Genome Biol. 2018;19(1). doi:10.1186/s13059-017-1382-0.

S24. Lake BB, Melo Ferreira R, Hansen J, et al. Cellular and Spatial Drivers of Unresolved Injury and Functional Decline in the Human Kidney. bioRxiv. 2025. doi:10.1101/2025.09.26.678707.

S25. Puthumana J, Thiessen-Philbrook H, Xu L, et al. Biomarkers of inflammation and repair in kidney disease progression. J Clin Invest. 2021;131(3). doi:10.1172/jci139927.
